# The Molecular Signature of Neuropathic Pain in a Human Model System

**DOI:** 10.1101/2024.01.04.23299847

**Authors:** Oliver P Sandy-Hindmarch, Pao-Sheng Chang, Paulina Simonne Scheuren, Iara De Schoenmacker, Michèle Hubli, Constantinou Loizou, Stephan Wirth, Devendra Mahadevan, Akira Wiberg, Dominic Furniss, Franziska Denk, Georgios Baskozos, Annina Schmid

## Abstract

Peripheral neuropathic pain remains challenging to treat, partly due to our limited understanding of the molecular mechanisms at play in humans. In this multicentre cohort study, we describe the local molecular signature of neuropathic pain at the lesion site, using peripheral nerves of patients with Morton’s neuroma as a human model system of neuropathic pain.

Plantar tibial nerves were collected from 22 patients with Morton’s neuroma (18 female, median age 60.0 [IQR 16.0] years) and control nerves from 11 participants (4 females, 58.0 [21.0] years) without a nerve injury. Pre-surgery, we collected data on pain severity, duration and nature (e.g., neuropathic pain inventory, NPSI).

RNA bulk sequencing of peripheral nerves identified 3349 genes to be differentially expressed between Morton’s neuroma and controls. Gene ontology enrichment analysis and weighted gene co-expression network analyses (WGCNA) revealed modules specific for host defence and neurogenesis. Deconvolution analysis confirmed that the densities of macrophages as well as B-cells were higher in Morton’s neuroma than control samples. The findings for T-cells were inconclusive. Modules associated with defence response, neurogenesis and muscle system development correlated with paroxysmal and evoked pain in people with Morton’s neuroma. Macrophage cell populations identified by deconvolution analysis as well as single differentially expressed genes (*MARCO, CD163, STAB1;* indicating the presence of a specific M(GC) subset of macrophages) correlated with paroxysmal pain.

Immunofluorescent analyses confirmed the presence of demyelination, higher densities of intraneural T-cells and CD163^+^MARCO^+^ macrophage subsets in Morton’s neuroma compared to control nerves. Histological CD68^+^ macrophage density correlated with burning pain. Our findings provide detailed insight into the local molecular signature in the context of human focal nerve injury. There is clear evidence for an ongoing role of the immune system in chronic peripheral neuropathic pain in humans, with macrophages and specifically the M(GC) MARCO^+^ subset implicated.

## Introduction

Neuropathic pain is a debilitating condition that affects ∼7% of the population.^1^ Compared to pain of nociceptive origin (e.g., musckuloskeletal pain), neuropathic pain is associated with higher pain severity^2^, lower quality of life^3^ and higher health care costs.^2^ The most common cause of neuropathic pain are peripheral neuropathies, and in particular entrapment neuropathies.^4^ Peripheral neuropathic pain remains challenging to treat^3^ with currently available pharmacological options providing limited benefit, often with significant side effects.^5,6^ These inadequacies of treatments stem in part from a lack of understanding of the pathophysiology of peripheral neuropathic pain and the limited translation of findings from preclinical models to humans.^7,8^ Calls to use human tissues to understand disease pathology are thus growing.^9,10^

However, access to human peripheral nerve tissues, however, remains a major challenge, especially in the context of neuropathic pain. There is a growing body of evidence of dorsal root ganglia molecular signatures associated with neuropathic pain caused by tumour lesions,^11,12^ providing important insights into changes in gene expression at the level of the sensory cell bodies. These studies found a dominant role of neuroimmune mechanisms in the context of neuropathic pain. Only few studies have investigated molecular changes at the sites of nerve injury in the distal nerve trunk. While some studies have focussed on a few selected genes using PCR methodology, ^13^ two studies have used bulk RNA sequencing to characterise the transcriptomic signature in human distal nerve trunks. Ray *et al.*^14^ used post-mortem tibial nerve tissues and identified sexually dimorphic expression of genes involved in pain, inflammation and neuro-immunity. Welleford *et al.*^15^ used serial sural nerve biopsies to identify gene signatures associated with acute nerve repair following transection injury. They identified genes representing antiapoptotic signalling, neurotrophic factor processes, cell motility and again immune cell chemotactic signalling to be differentially expressed. In both studies though, no clinical information was available on the presence or nature of neuropathic pain, thus precluding any inferences about the local molecular signature associated with the clinical features of neuropathic pain.

Here, we used Morton’s neuroma as a unique model system to study the local molecular signature at the site of nerve injury in the context of neuropathic pain. Despite its misleading name, Morton’s neuroma is a peripheral entrapment neuropathy, in which a plantar digital nerve is compressed just proximal to its bifurcation under the transverse intermetatarsal ligaments of the foot.^16^ Patients develop typical neuropathic symptoms, including burning, paroxysmal pain and paraesthesia.^17^ In some patients, treatment involves the surgical excision of the affected nerve, therefore offering a rare and unparalleled source of injured human tissue in the context of neuropathic pain. Using clinical phenotyping of neuropathic pain followed by transcriptomic analyses of Morton’s neuroma compared to human control nerves, we aimed to describe the molecular signature of neuropathic pain and validate our results using immunohistochemistry. Our findings indicate gene signatures related to neurogenesis and the immune system. Macrophage populations identified by deconvolution analysis and a differentially expressed gene signature, characteristic of a specific M(GC) macrophage subset (*CD163+, MARCO+, STAB1+*), correlated with the severity of paroxysmal pain. Histological validation confirmed demyelination as well as increased intraneural T cell and CD163+MARCO+ macrophage densities persisting in the chronic stages of Morton’s neuroma.

## Material and methods

### Study design and participants

This study reports cross sectional data from a prospective longitudinal multicentre cohort study. Twenty-two adults over the age of 18 years with clinically diagnosed Morton’s neuroma who were listed for neuroma excision surgery were recruited from waiting lists of Oxford University Hospitals and Royal Berkshire NHS Foundation Trusts UK (n=13) or Balgrist University Hospital, Zurich, Switzerland (n=9) between 13/3/2019 and 22/06/2021 (interrupted by the COVID-19 pandemic). Patients were excluded if they had systemic diseases affecting neural tissues (e.g., diabetic neuropathy), other neurological conditions affecting the lower limb (e.g., lumbar radiculopathy), severe somatic diseases affecting the foot (e.g., polyarthritis), previous extensive surgery of the forefoot which may have an impact on sensory function, cortisone injection for the operated Morton’s neuroma within the preceding 3 months, or if they were pregnant. We also recruited eleven control participants from Oxford University Hospitals NHS Foundation Trust who underwent surgery in which unaffected neural tissue could be sampled. These included surgeries for nerve grafting, muscle flap or lower limb amputations (e.g., due to non-healing fracture). Control participants did not have any current systemic or neurological disease and no clinical history of neuropathy affecting the excised control nerve. Ethical approval was received from the South Central - Oxford C Research Ethics Committee (REF 18/SC/0410), the London - Camden & Kings Cross Research Ethics Committee (REC16/LO/1920) and the cantonal ethics committee Zurich (2018-02198). All participants provided informed written consent in accordance with the declaration of Helsinki.

Whereas control participants provided demographic and medical history data on the day of surgery, patients with Morton’s neuroma attended a pre-operative session with an investigator.

### Phenotypic data of the Morton’s neuroma cohort

Age, sex, BMI and duration of Morton’s neuroma symptoms (if applicable) were recorded for each participant. The severity of patients’ symptoms was evaluated pre-surgery using a range of clinical questionnaires. Neuropathic pain severity was evaluated with the Neuropathic Pain Symptom Inventory (NPSI),^18^ which is formed of 10 numerical rating scales from 0 (no pain) to 10 (worst pain imaginable). These scales are used to rate pain subtypes including burning, deep pressure pain, paraesthesia, paroxysmal symptoms and evoked pain, as well as a composite score. The average severity of pain and numbness over the past 24 hours were recorded on separate visual analogue scales (VAS), where 0 indicated no symptoms, and 10 indicated the worst symptoms imaginable.

### Tissue collection and preparation

From patients with Morton’s neuroma, 22 affected plantar digital nerves were collected during routine surgical excision of the neuroma. Each Morton’s neuroma sample was split into four pieces, two from the site of the neuroma just proximal to the bifurcation of the plantar digital nerve, and two just distal to the bifurcation (digital branches, Supplemental Figure 1). One distal and one proximal sample were placed into RNAlater® solution (ThermoFisher Scientific, AM7021) and stored overnight at 4°C. The remaining distal and proximal samples were placed in 4% paraformaldehyde solution and stored at room temperature overnight. Only the proximal samples representing the site of the Morton’s neuroma were used in our analyses.

From control participants, 11 nerves of the upper and lower limbs were collected (five nerve grafts, three from muscle flaps, three from amputations). Of these 11 control samples, three were plantar digital nerves, three were gracilis motor nerves, four were posterior interosseous sensory nerves and one was an intercostal sensory nerve. Each sample was split into two samples and placed into RNAlater® and paraformaldehyde as detailed above.

For molecular analyses, the proximal nerve samples in RNAlater® solution had any extraneural connective tissue removed the next day. The samples were then snap frozen in an Eppendorf tube in liquid nitrogen and stored at -80 °C for batch RNA extraction. On the day of RNA extraction, 1mL of TRIzol^TM^ solution (ThermoFisher Scientific, 15596026) was added to the frozen sample which was then homogenised. Chloroform was added before centrifugation for phase separation. Total RNA extraction was done using a hybrid method of phenol extraction (TriPure; Roche, Welwyn Garden City, United Kingdom) combined with column purification (High Pure RNA tissue Kit; Roche). The aqueous liquid phase containing the nucleic acids was removed and added to the columns of the High Pure RNA tissue kit (Roche Diagnostics). RNA was purified using repeated wash steps and DNAse treatment. The concentration of RNA in the samples was measured using a nanodrop. Total RNA was provided to the sequencing centre, and the poly-adenylated fraction was selected for sequencing.

For histological analyses, the samples stored overnight in paraformaldehyde solution were washed 3 times in 0.1M phosphate buffer and stored in a 20% w/v sucrose solution for three days at 4°C. Nerve samples were frozen in optimal cutting temperature gel in base moulds and frozen in liquid nitrogen. Samples were then stored at -80°C for immunofluorescence analyses.

### RNA sequencing

RNAseq was performed at the Wellcome Centre for Human Genetics. All samples passed initial QC assessing RNA degradation (RIN >8). Polyadenylated transcript enrichment and strand specific library preparation was done using the NEBNext^®^ Ultra^TM^ II RNA Library Prep kit (NEB, E7770) following manufacturer’s instructions. Libraries were amplified using unique dual indexing primers, based on Lamble *et al.*^19^ Paired end sequencing was performed on an Illumina NovaSeq6000 system.

Quality control of the raw sequencing reads was performed using FastQC/ MultiQC and assessed the yield, number and percentage of reads duplicates, the per sequence Phred quality score, the GC content, the length distribution and over-representing sequences or adapter contaminations. Two of the sequenced libraries had an over-represented sequence that indicated adapter contamination (1.38% and 3.27%). These were dealt by utilising the “soft clipping” technique of aligner.

Reads were mapped to the GRCh38 human genome by the STAR aligner programme^20^ with standard ENCODE options, and gene counts were determined using HTSeq.^21^ The fraction of uniquely mapped reads across samples was excellent, median = 94.13% (IQR 93.45% - 94.52%), total number of pairs of reads median = 18953,721 (IQR 18165,457 – 20463,241). DESeq2^22^ was used to determine differential gene expression between nerve samples from patients with Morton’s neuroma and control participants, controlling for gender effects by using an additive design of ∼ gender + condition. Moderation of log fold changes was carried out by using a zero-mean normal prior on the coefficients of interest. Gene ontology (GO) analyses were performed in R using the GOseq^23^ package. A probability weighting function was used to model gene length bias for the GO analysis, and the background consisted of all expressed genes with more than 20 counts in at least 50% of the samples. Genes with an FDR adjusted p-value < 0.05 and in the top 25% (3^rd^ quantile) of log 2-fold changes (absolute LFC > 0.64) were considered significantly regulated for this and all downstream analyses. Unsupervised medoid - “pam” clustering was used to identify groups of genes and samples with similar expression profile. The optimal number of clusters was determined using the gap statistic. Hierarchical clustering was carried out using Ward’s method.

We then performed Weighted Gene Co-expression Network Analyses (WGCNA) using the WGCNA^24^ package in R. WGCNA is a method that builds a co-expression network based on the correlation between different genes and then identifies modules of highly correlated genes. An unsigned co-expression network was built using the top 25% of expressed genes (20 counts in at least 50% of samples) ranked by observed variance, and modules were identified using the dynamic tree cut algorithm.^8^ We assigned biological functions to these modules by looking at over-represented GO biological process terms. We then used the module eigengene, i.e. the 1^st^ principal component of a given module, as the representative gene for each module to calculate associations with phenotypic traits. Deconvolution was done using the deconvolution pipelines from EPIC,^25^ quanTIseq^26^ and GEDIT^27^ software packages and online tools. In the absence of reference sequences from human peripheral nerves, additional sequences were used from the ABIS^28^ and LM22^29^ datasets.

### Immunofluorescent analyses

Immunofluorescent staining was done on proximal nerve samples from patients with Morton’s neuroma and control participants to detect intraneural macrophages and T-cells and their specific subtypes. We also stained for the neuronal markers myelin basic protein (MBP) and β-tubulin to determine the extent of axonal loss and demyelination. For each participant, 14µm serial tissue sections were cut using a cryostat (Leica CM1860 UV, Germany) and adhered to SuperFrost Plus^TM^ microscope slides (VWR, 631-0108) coated with 0.01% poly-L lysine solution (Sigma Aldrich, P8920). 400µl of 0.01% PLL solution was added to each slide and left to dry at room temperature. The tissue sections were left to dry overnight at room temperature before being stored at -20°C for batch staining.

Before staining, the tissue sections were defrosted at room temperature for an hour and incubated at 60°C for another hour to improve tissue adherence. Then, mild heat-induced epitope retrieval (HIER) was applied for 5 hours by incubating the sections in 70°C Tris-EDTA buffer (pH 9.0) containing 10mM Trisma Base (Sigma-Aldrich, T6066), 1mM EDTA (Sigma-Aldrich, ED2SS) and 0.05% Tween-20 (Sigma-Aldrich, P7949) in distilled water. After HIER, the sections were washed once for 5 minutes in PBS-Tx: 0.2% Triton X-100 (Sigma-Aldrich, X100) in PBS. Then, the sections were incubated for an hour at room temperature in blocking solution containing 5% normal goat serum (NGS, Sigma-Aldrich, G6767), 1% bovine serum albumin (BSA, Europa Bioproducts Ltd., EQBAH65), 1% dimethyl sulfoxide (DMSO, Sigma-Aldrich, D5879), 0.5% skim milk powder (Sigma-Aldrich, 70166), 0.3% Triton X-100 and 0.1% NaN_3_ (Sigma-Aldrich, S8032) in PBS.

For evaluating the integrity of axons (β-tubulin III) and myelin sheaths (myelin basic protein, MBP) and the density of immune cells of interest (CD68, CD3, CD4, MARCO, CD163), we double stained with the markers summarised in Supplemental Table 1. The tissue sections were incubated with primary antibodies overnight at 4°C in a humidified chamber. The next day, sections were washed five times with PBS-Tx. Sections were then incubated with secondary antibodies for two hours at room temperature in a humidified chamber. Lastly, the tissue sections were washed four times with PBS-Tx and once with PBS before mounted in Vectashield^®^ mounting media with DAPI (Vector Laboratories, H-1200). For CD163/MARCO staining, Nuclear Violet (AAT Bioquest, 17543, 1:1000) was used to visualise nuclei.

The stained sections were imaged with an Observer Z1 confocal imaging system (Zeiss, Germany) at 200-fold magnification, and maximum intensity projections were derived from Z-stacks. Depending on tissue area, 1-7 images (average 3) were taken per tissue sample for quantification purposes. The density of CD3^+^, CD68^+^, CD3^+^CD4^+^, CD163^+^, MARCO^+^, CD163^+^MARCO^+^ and CD163^+^MARCO^-^ cells and the amount of β-tubulin III and MBP were quantified by an examiner blinded to group allocation using ImageJ (version 2.9.0). For β-tubulin III and MBP, intraneural areas were selected and corrected mean grey values were determined by subtracting background mean grey values (average of three extra-neural areas) from intraneural values. Area fractions were quantified as the percentage of intraneural pixels with positive signal (threshold = 30). Intraneural CD3^+^, CD68^+^, CD3^+^CD4^+^, MARCO^+^, CD163^+^, CD163^+^MARCO^+^ and CD163^+^MARCO^-^ cells were counted using the multipoint tool, and densities established as cells/mm^3^. Only cells containing a DAPI/Nuclear Violet positive nucleus were counted.

### Statistics

Sample size is based on a simulation of gene counts from our previous human RNAseq experiment in skin of patients with and without nerve injury and neuropathic pain.^30^ A sample of n=11-18 will have 69-80% power to detect 10% differentially expressed genes assuming log normal distribution of fold changes with mean log_2_1.5, a standard deviation 0.5*log_2_1.5 and a sequencing depth of 24M 75bp paired-end.

The statistical and graphical analyses were conducted using R,^31^ SPSS (version 28, IBM) and Prism 9 (version 9.5.1, GraphPad). Visual inspection as well as Shapiro-Wilk tests were used to test for normality within the data. The Wald test was used to determine differences in gene expression with an FDR-adjusted p-value < 0.05 and an absolute log2 fold change > 0.64 deemed significant. This set of significantly regulated genes was considered for the GO enrichment analysis, which was performed separately for up- and downregulated genes. To determine the differences between deconvolution-derived cellular proportions, independent sample Mann-Whitney U tests were applied with an FDR-adjusted p-value < 0.05 deemed significant.

For comparing the integrity of β-tubulin III and MBP as well as intraneural immune cell density between groups, independent sample Mann-Whitney U tests or Student’s t-tests were applied with p-values < 0.05 deemed significant.

The correlations between WGCNA modules, their single genes or immunohistological data and patients’ pain-related symptoms (NPSI subscores and VAS for numbness or pain) were evaluated using the Spearman’s rank correlation or Pearson’s correlation with an FDR-adjusted p-value < 0.05 considered significant.

### Data availability

The RNA sequencing data that support the findings of this study are openly available in GEO following publication in a peer reviewed journal (series number GSE250152). Data on clinical phenotypes are available from the corresponding author upon reasonable request.

## Results

### Demographic and clinical data of participants

Demographic and clinical data for study participants are shown in Table 1. Age, height, weight and BMI were comparable, but the percentage of female participants was higher in the Morton’s neuroma compared to the control group (82 vs 36%). Patients had on average a moderate severity of pain (median VAS 4.46 [IQR 4.42]).

**Table 1:** Demographic and clinical data. Data are presented as median [interquartile range] unless stated otherwise.

|  | Control | Morton's Neuroma |
| --- | --- | --- |
| Number of Participants | 11 | 22 |
| Age (years) | 58.0 [21.0] | 60.0 [16.0] |
| Female gender, n (%) | 4 (36.4%) | 18 (81.8%) |
| Height (cm) | 177.0 [18.0] | 167.5 [12.0]<sup>^</sup> |
| Weight (kg) | 78.0 [46.0] | 70.0 [21.8]<sup>^</sup> |
| BMI (kg/m<sup>2</sup>) | 24.9 [10.6] | 24.9 [6.3]<sup>^</sup> |
| Duration of symptoms (months) |  | 30.0 [18.0]<sup>^</sup> |
| NPSI total score |  | 30.0 [18.0]<sup>^</sup> |
| burning pain |  | 5.0 [8.0]<sup>^</sup> |
| deep pain |  | 3.0 [3.0]<sup>^</sup> |
| evoked pain |  | 3.5 [2.5]<sup>^</sup> |
| paroxysmal pain |  | 5.0 [7.0]<sup>^</sup> |
| paraesthesia |  | 2.0 [5.3]<sup>^</sup> |
| Visual Analogue Scale (average over past 24 hours) |  |  |
| Pain |  | 4.5 [4.4]<sup>^</sup> |
| Numbness |  | 0.6 [4.3]<sup>^</sup> |

<sup>^</sup> Clinical data from one person with Morton's neuroma missing as questionnaires not completed.

### Differential gene expression analysis reveals a high level of differentially expressed genes

To determine a molecular signature associated with neuropathic pain, differential gene expression analysis was conducted on the 22 plantar digital nerves from people with Morton’s neuroma and the 11 control nerves. A Principal Component Analysis (PCA) plot (Figure 1A) shows good separation between the patient and control nerve samples. The majority of the variance can be attributed to Principal Component 1 (PC1), which accounts for 46% of variance within the data, PC2 accounts for a further 13% of the variance.

**Figure 1:**
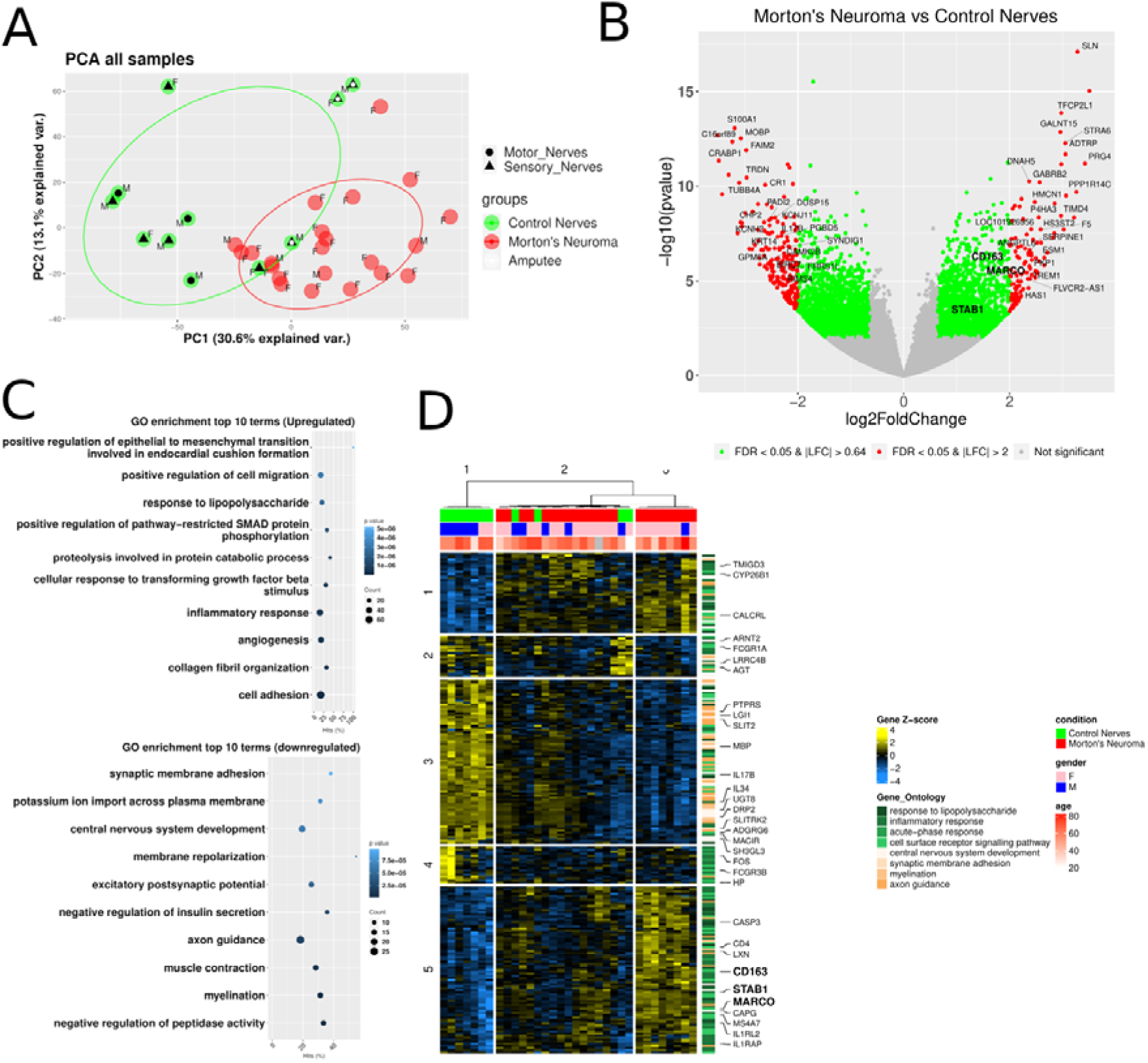
Differential gene expression analysis revealed a high number of differentially expressed genes between samples from patients with Morton’s neuroma and control participants. **(A)** Principal component analysis plot showing good separation between patients with Morton’s neuroma (red) and control participants (green). Control nerves from amputations are denoted by a blue dot; control motor and sensory nerves are denoted by a black circle or triangle respectively. Circles represent the centroids and 95% CI of the respective groups. The first principal component accounted for 30.6% of the variance within the data and the second principal component accounted for 13.1%. Gender of participants is indicated with M – Males and F – Females. **(B)** Volcano plot showing the level of differential gene expression. Genes shaded in red have an FDR adjusted p-value < 0.05 and a an absolute Log2FC > 2. Genes shaded in green have an FDR adjusted p-value < 0.05 and an absolute Log2FC > 0.64, the top 25% quantile. Genes shaded in grey are non-differentially expressed genes. **(C)** Dot plot of the top 15 enriched gene ontology terms ranked by their over-representation p-value for both up and down regulated genes in Morton’s neuroma vs controls. Hits (%) indicates the fraction of genes in the term that are differentially expressed. Counts indicate the number of genes in the term that are differentially expressed. **(D)** Heatmap showing the clustering of samples based on the expression levels of genes that belong in the top enriched biological processes associated with the response of the immune system and the development of the nervous system.

Differential gene expression analysis identified over 3349 genes to be differentially expressed (DEG, Figure 1B), 1461 upregulated and 1888 were downregulated. A large proportion of these genes had an absolute Log2 fold change (FC) of above 2. A GO enrichment analysis for the biological processes over-represented showed that amongst genes upregulated in Morton’s vs control nerves, there were significantly over-represented terms related to the response of the immune system to inflammation including “response to lipopolysaccharide” and “inflammatory response”, Figure 1C. Amongst downregulated genes there was a significant enrichment of processes related to the development of the nervous system including “axon guidance”, “myelination”, “synaptic membrane adhesion”, “central nervous system development”, Figure 1C. A heatmap of the 238 differentially expressed genes that were annotated with the selected highly enriched terms related to the immune system and development of the nervous system (Figure 1D), showed a mostly neuronal cluster of genes (Cluster 3) downregulated in Morton’s neuroma, two mixed clusters (Cluster 1 and 2) with both inflammatory response and nervous system development related terms. A small cluster of immune response related genes (Cluster 4) was downregulated in Morton’s neuroma, but the main cluster of inflammatory related genes (Cluster 5) was consistently upregulated in Morton’s neuroma vs control nerves.

### Immune and defence response are over-represented pathways

A Weighted Gene Co-Expression Network Analysis (WGCNA) was conducted to identify groups / modules of genes with similar expression patterns. We then determined over-represented biological processes within these modules and looked for associations between the modules’ representative genes (eigengene) and phenotypic traits.

In the WGCNA analysis, we identified nine modules (named after the top over-represented GO term amongst the genes belonging in the module), that were enriched for the biological processes of immune and defence responses, regulation of neuron death and neurogenesis, collagen metabolism, muscle systems, carbohydrate transmembrane transport, mitochondrion organisation and reproductive behaviour. The eigengene of the module associated with defence response was positively correlated (r = 0.66 95%CI [0.32, 0.85], p-value = 0.0007, FDR = 0.004) and the module eigengene associated with neurogenesis was negatively correlated (r = -0.54 95%CI [-0.14, -0.79], p-value = 0.009, FDR = 0.05) with NPSI paroxysmal pain. The module eigengene associated with muscle system process was negatively correlated (r = -0.44 95%CI [-0.01, -0.73], p-value = 0.03, FDR = 0.2) with NPSI evoked pain (Figure 2A). These were moderate to strong correlations that all reached nominal significance, while the correlation of the defence response module eigengene with NPSI paroxysmal pain survived FDR adjustment. We then looked at the individual expression patterns of the top 25% genes belonging to these modules ranked by their correlation strength with the module eigengenes. We observed one group of Morton’s neuroma patients with decreased expression in the defence response genes and lower paroxysmal pain; one group with higher expression in defence response genes and lower expression of muscle system process genes with higher paroxysmal and lower evoked pain; and finally a third group with higher expression in neurogenesis genes and mixed expression of muscle process and defence response genes with a distinct anti-correlated expression pattern in the paroxysmal and evoked pain. Unsupervised clustering of genes revealed two clusters: Cluster 1 had the genes in the modules associated with defence response and neurogenesis and Cluster 2 the genes associated with muscle system process (Figure 2B, Supplemental File 1).

**Figure 2:**
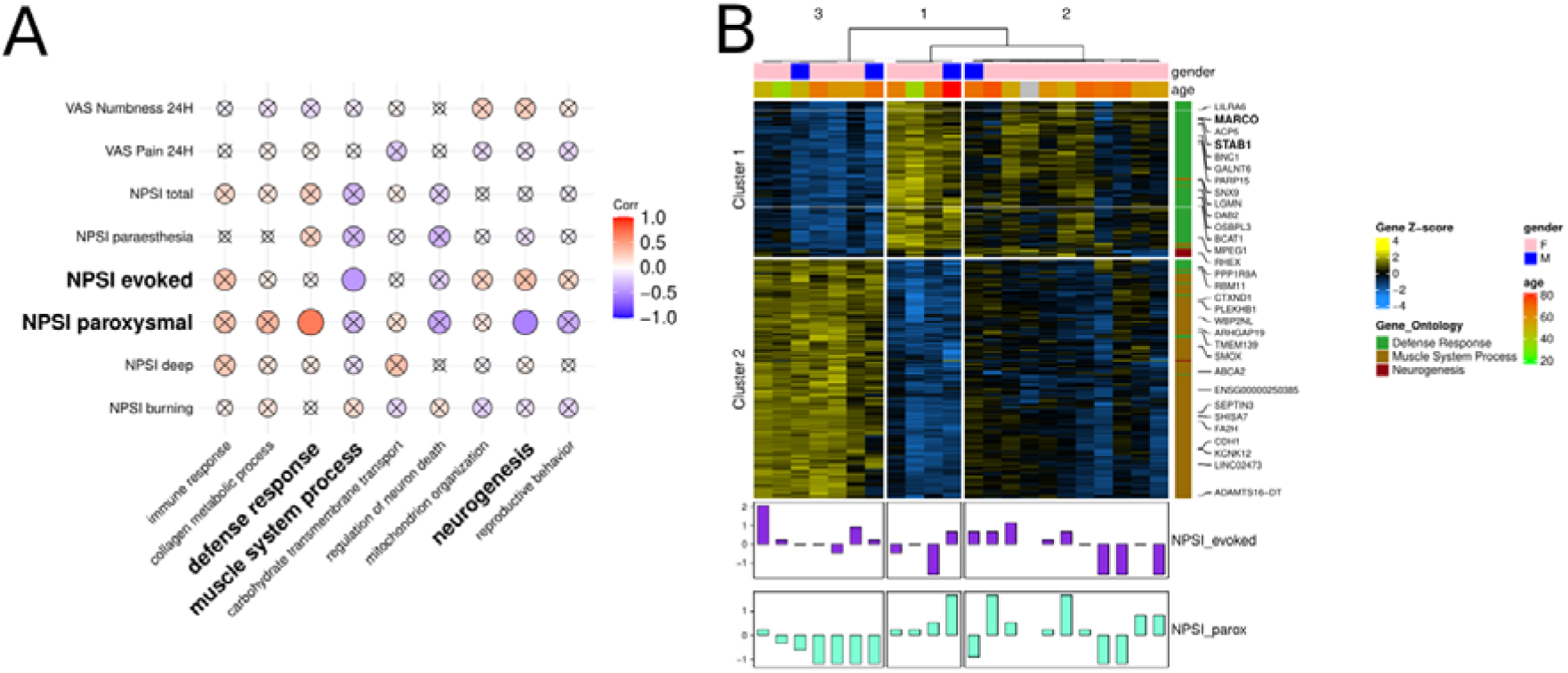
Weighted Gene Co-Expression Network analysis. **(A)** Correlation plot showing the Pearson’s correlation strength between phenotypic traits associated with the symptoms and modalities of pain and the eigengenes of the modules identified through the co-expression network analysis. The eigengenes are named after the top enriched biological process in the module. Correlation strength is encoded in the size and colour of the dots. Non nominally significant associations (p-value > 0.05) are x crossed. **(B)** Heatmap showing the expression patterns and unsupervised clustering of samples based on the genes that belong in the modules that had significant correlations with phenotypic traits. The significantly correlated symptoms are z-transformed and shown in barplots.

### Deconvolution analysis identifies significant differences in immune cell populations

The large number of DEGs between Morton’s and control nerves likely represents a change in, or recruitment of cell types, such as during an inflammatory or fibrotic response. Using deconvolution analysis to quantify immune cell proportions, we identified significant differences between samples of patients with Morton’s neuroma and controls. Proportions of immune cell types including macrophages and Memory B cells were higher in Morton’s neuroma compared to control samples (Figure 3A-C). The packages we used to deconvolute used the traditional division of macrophages into M1 and M2, rather than updated nomenclature.^32^ Both types were increased in the neuroma, with the full list of genes denoted as ‘M1’ and ‘M2’ in the package provided in Supplemental Table 2. CD4^+^ helper T-cells were lower in patients with Morton’s neuroma (Figure 3D). Both M1 and M2 macrophage cell proportions correlated with the severity of paroxysmal pain (Figure 3E-F).

**Figure 3:**
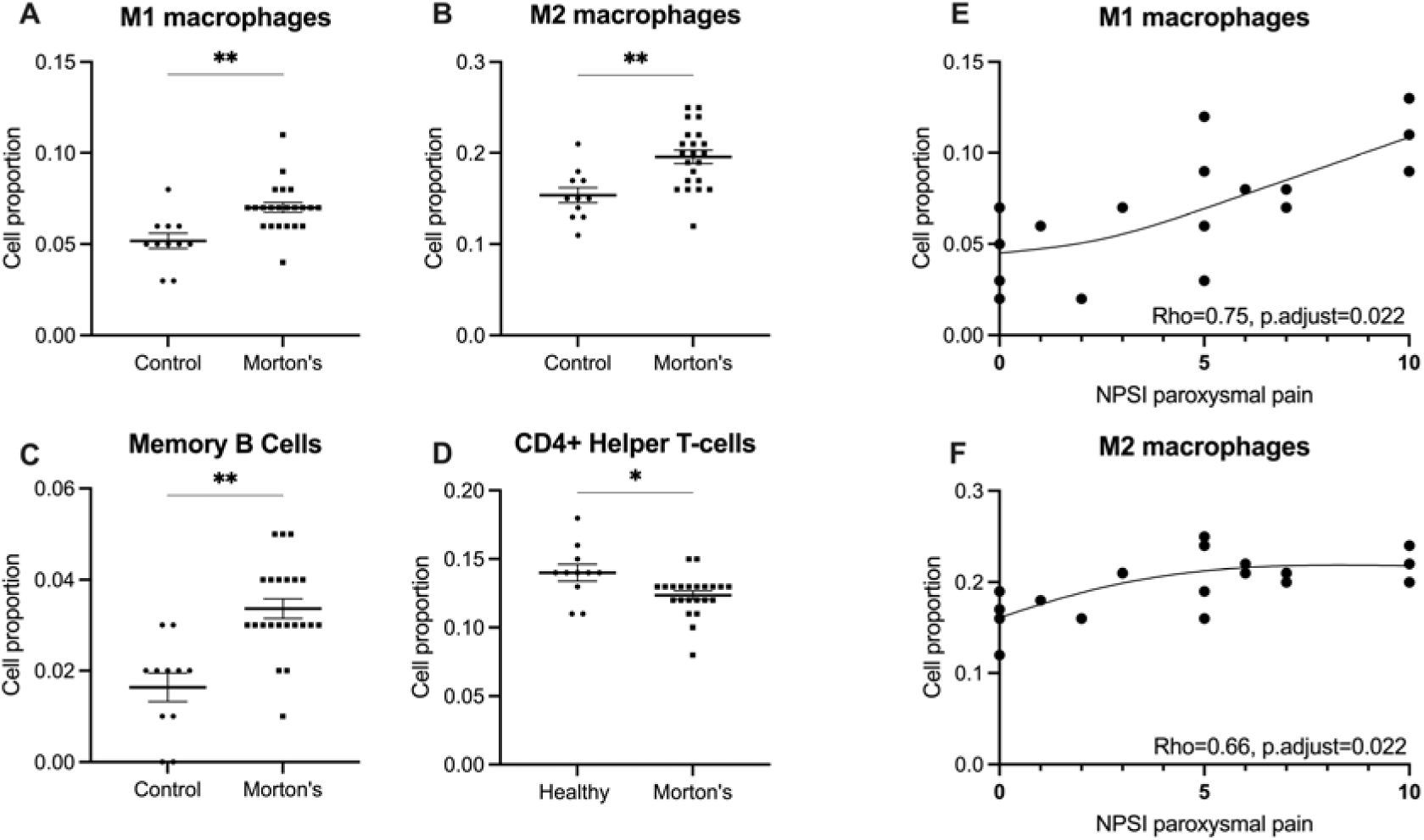
Deconvolution analysis reveals significant differences in immune cell proportions between samples from patients with Morton’s neuroma and controls. **(A-C)** Macrophages and memory B-cell proportions were found to be significantly higher in samples from patients with Morton’s neuroma compared to control participants. The package we used distinguished between macrophages according to the traditional M1 and M2 nomenclature. **(D)** CD4^+^ helper T-cell proportions were found to be significantly decreased in Morton’s neuroma compared to control samples. **(E,F)** Both M1 and M2 macrophage proportions significantly correlate with NPSI paroxysmal pain. Significant differences between groups were determined by FDR corrected Mann-Witney U tests (adjusted p <0.05). Spearman’s rank correlation was used with an FDR adjusted p<0.05. NPSI = Neuropathic pain symptom inventory.

### Genes specific to M(GC) MARCO^+^ subset of macrophages are upregulated and correlate with clinical phenotypes

Given the consistent signature for immune cells and specifically the association of macrophage signatures with paroxysmal pain, we further screened our list of DEGs, specifically those in co-expression modules that correlated with symptoms in participants with Morton’s neuroma, or were annotated with significantly enriched GO terms associated with the immune response and the development of the nervous system. Amongst these genes, the following combination stood out as being known markers of a specific macrophage phenotype according to updated nomenclature:^32^ (1) *MARCO* - Macrophage Receptor With Collagenous Structure; (2) *CD163* – typically found in resident macrophages and a high-affinity scavenger receptor for the haemoglobin-haptoglobin complex; (3) *STAB1*: Stabilin-1, a transmembrane receptor protein that is involved in angiogenesis, cell adhesion and receptor scavenging. These three genes are signature markers for M(GC) macrophages, i.e. those that have been stimulated with glucocorticoids.^32^ We looked for associations between these genes and symptoms in participants with Morton’s neuroma. All three genes were positively correlated with NPSI paroxysmal pain and paraesthesia (Figure 4A, B). *MARCO* had a strong and nominally significant correlation with paroxysmal pain (r = 0.48 95% CI [0.1, 0.76], p-value = 0.02). All correlations are shown in Supplemental Table 3.

**Figure 4:**
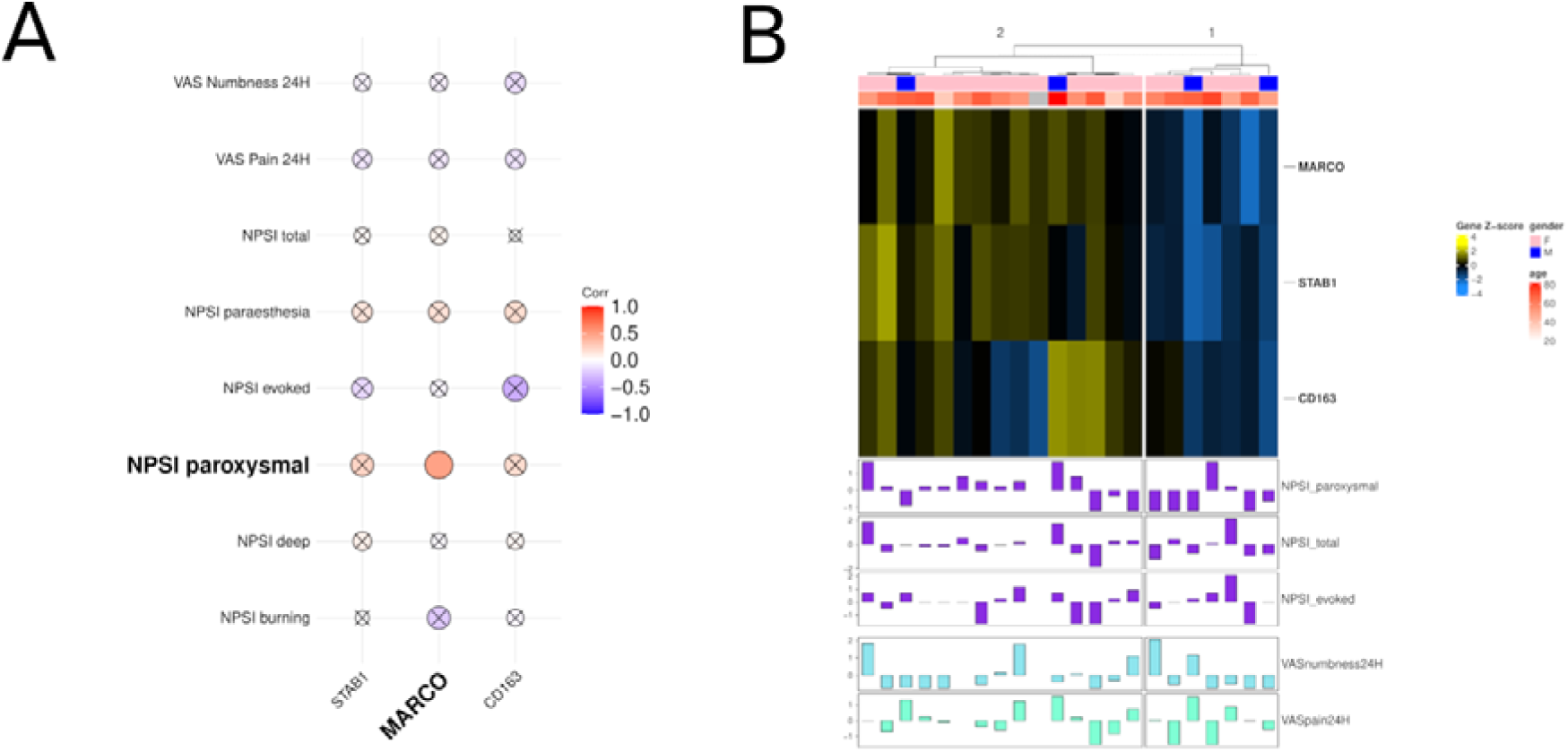
Individual genes representing a M(GC) MARCO^+^ subset of macrophages correlated with clinical pain phenotypes. **(A)** The expression of individual genes *MARCO, STAB1, CD163*, denoting M(GC) macrophages, are correlated with clinical pain phenotypes. *MARCO* is strongly and nominally significantly correlated with paroxysmal pain. **(B)** Heatmap showing the normalised expression of these genes in Morton’s neuroma participants. A cluster of participants with increased expression of *MARCO* also has increased NPSI paroxysmal pain.

### Immunofluorescence confirms demyelination and M(GC) MARCO^+^ macrophage infiltration in Morton’s neuroma

To validate the contribution of myelination in the GO analysis, we used immunofluorescent analyses to determine the integrity of axons and myelin sheaths. We confirmed a reduction of the corrected mean grey values and area fractions of MBP in nerves from patients with Morton’s neuroma compared to control nerves, indicative of demyelination (Figure 5A-C).

**Figure 5:**
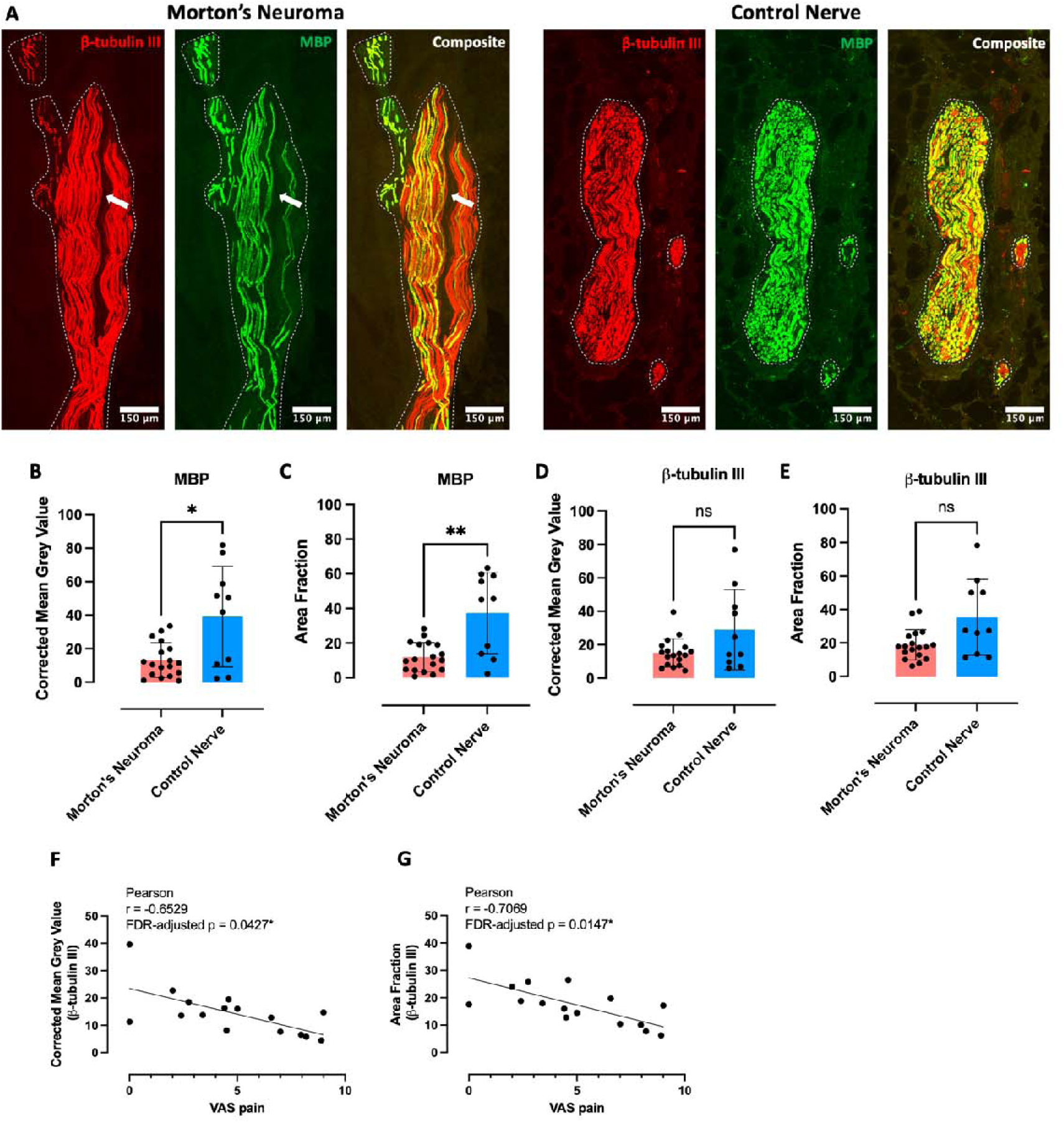
Immunofluorescence analysis confirms demyelination in Morton’s neuroma. **(A)** Representative images of nerve sections from a patient with Morton’s neuroma (left) and a healthy individual (right) stained with anti-β-tubulin III (red) and MBP (green). The boundaries of intraneural areas are indicated with white dotted lines. We repetitively identified several areas that are devoid of axons within the Morton’s neuroma. One such region is identified in A with a white arrow. **(B-C)** Quantification confirmed a reduction of MBP corrected mean grey values **(B)** and area fractions **(C)** in Morton’s neuroma compared to control nerves. **(D-E)** Corrected mean grey values and area fractions of β-tubulin III were comparable between groups. Morton’s neuroma, N=18; Control Nerve, N=10. Data are presented as means and standard deviations. Student’s t tests, p-values = 0.024 (B), 0.008 (C), 0.106 (D) and 0.051 (E). **(F)** Corrected mean grey value and **(G)** area fraction of β-tubulin III staining strongly correlated with patient’s pain measured on a visual analogue scale (VAS). N=16 (2 patients had missing VAS pain scores), Pearson’s correlation, FDR-adjusted p-values p=0.0427 and 0.0147 respectively.

There was no significant difference in β-tubulin III between groups (Figure 5A, D-E). A negative correlation of β-tubulin III’s area fraction and corrected mean grey value with the VAS for pain survived correction for multiple comparisons between axonal or myelination parameters with patients’ pain phenotype (Figure 5F-G, Supplemental Table 4).

Validation of immune cell infiltration revealed no significant differences in the density of intraneural CD68^+^ cells between groups (Figure 6A-B). In contrast, there was a higher density of intraneural T-cells in patients with Morton’s neuroma compared to healthy controls (Figure 6A, C). The density of intraneural CD68^+^ cells significantly correlated with NPSI burning pain (Figure 6D, Supplemental Table 5).

**Figure 6:**
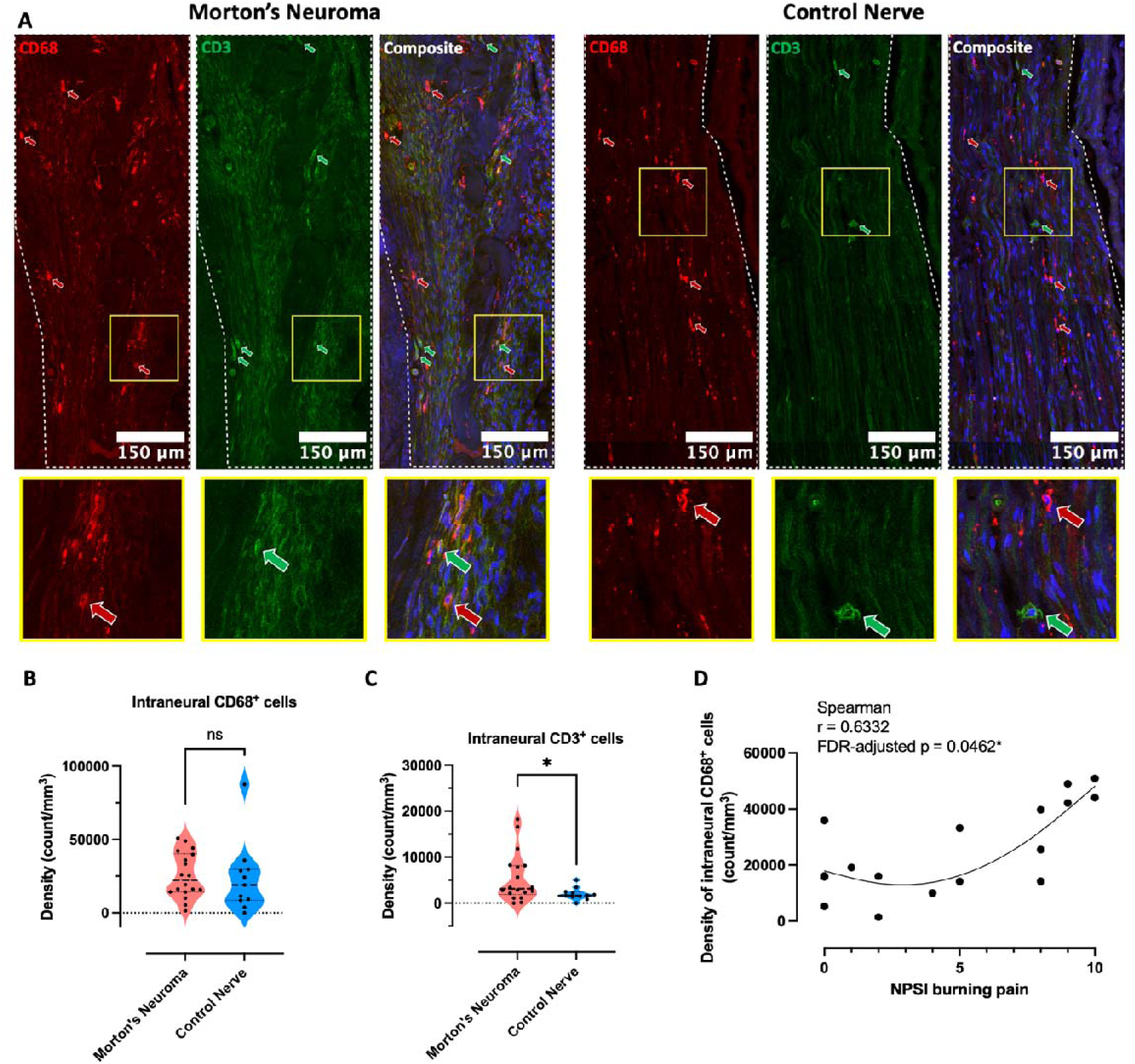
The density of intraneural CD3^+^ but not CD68^+^ cells is higher in patients with Morton’s neuroma. **(A)** Representative images of nerve sections from a patient with Morton’s neuroma (left) and a healthy individual (right) stained with CD68 (red) and CD3 (green). The boundaries of intraneural areas are indicated with white dotted lines. The areas within yellow squares were enlarged 7x and displayed below. The red and green arrows indicate examples of CD68^+^ and CD3^+^ cells respectively. **(B-C)** Unlike CD68^+^ cells **(B)**, the densities of intraneural CD3^+^ cells **(C)** were significantly higher in Morton’s neuroma compared to control nerves. Morton’s neuroma, N=18; Control Nerve, N=11. Data are presented as medians and interquartile ranges. Mann-Whitney U tests, p-values = 0.3397 and 0.0402, respectively. **(D)** The density of intraneural CD68^+^ cells was correlated with patients’ burning pain measured on the Neuropathic Pain Symptom Inventory subscore, N=18, Spearman’s correlation, FDR-adjusted p=0.046.

Further macrophage sub-phenotyping focusing on the M(GC) subset using markers CD163 and MARCO revealed higher intraneural densities of CD163^+^, CD163^+^MARCO^+^ and CD163^+^MARCO^-^ cells in Morton’s neuroma compared to control nerves (Figure 7A-E). No significant correlation was found between the intraneural densities of CD163^+^, MARCO^+^, CD163^+^MARCO^+^, or CD163^+^MARCO^-^ macrophages and patients’ pain phenotype (Supplemental Table 6 and 7).

**Figure 7:**
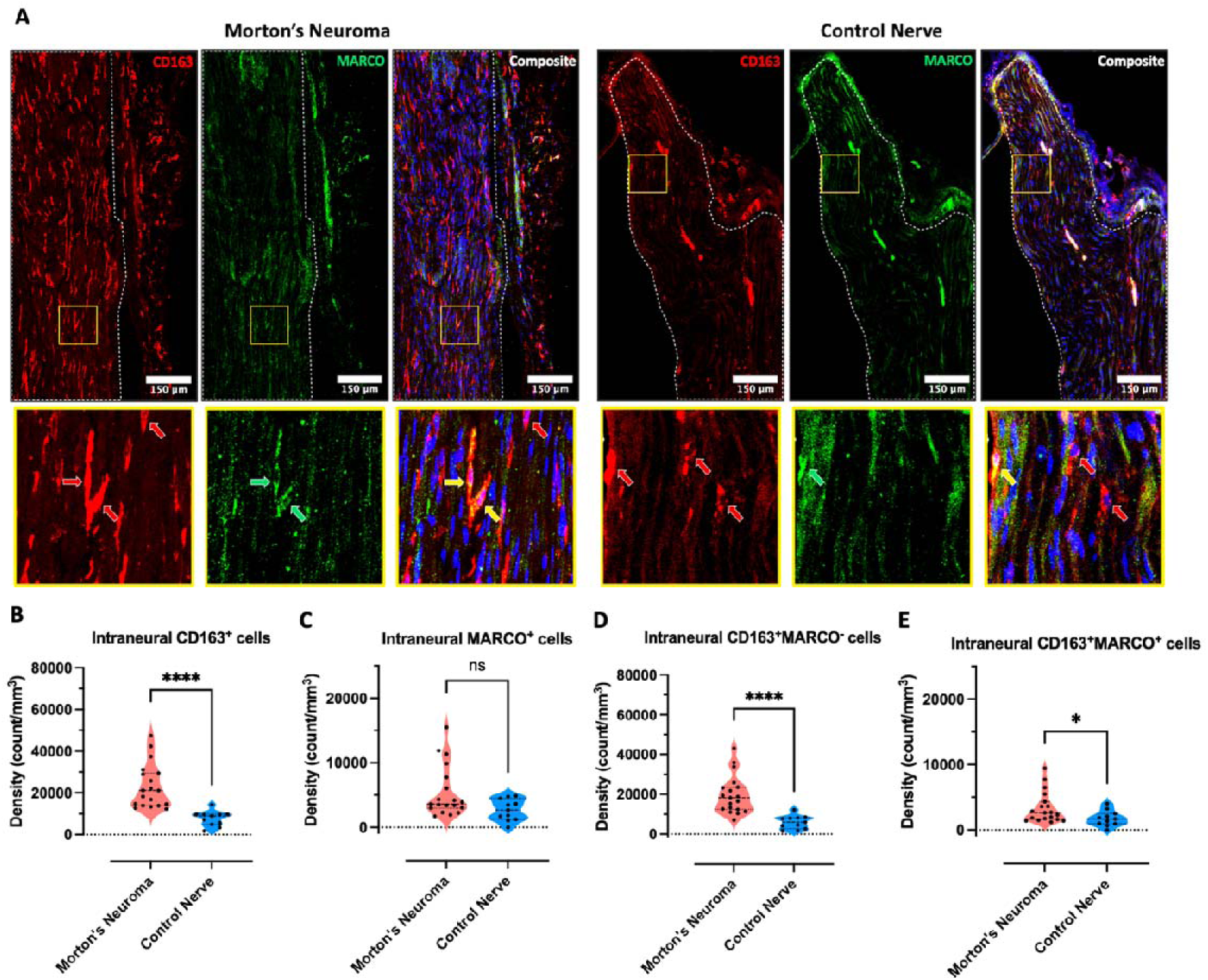
The densities of intraneural CD163^+^, CD163^+^MARCO^+^ and CD163^+^MARCO-macrophages are increased in Morton’s neuroma. **(A)** Representative images of nerve sections from a patient with Morton’s neuroma (left) and a healthy individual (right) stained with CD163 (red), MARCO (green) and Nuclear Violet (blue). The boundaries of intraneural areas are indicated with white dotted lines. The areas within yellow squares were enlarged 21.7x and displayed below. The red, green and yellow arrows indicate examples of CD163^+^, MARCO^+^ and CD163^+^MARCO^+^ cells respectively. **(B-E)** Except for intraneural MARCO^+^ cells **(C)**, the densities of intraneural CD163^+^ **(B)**, CD163^+^MARCO^+^ **(D)** and CD163^+^MARCO^-^ cells **(E)** were significantly higher in Morton’s neuroma compared to control nerves. Morton’s neuroma, N=19; Control Nerve, N=11. Data are presented as medians and interquartile ranges. Mann-Whitney U tests, p-values = <0.0001 (B), 0.070 (C), <0.0001 (D) and 0.0374 (E).

Further T-cell subtyping did not identify a significant difference in intraneural CD3^+^CD4^+^ cell densities between Morton’s neuroma and control nerves (Supplemental Figure 2). There was no significant correlation between the density of intraneural CD3^+^CD4^+^ cells with patients’ pain phenotype (Supplemental Table 8). We did not identify any CD3^-^CD4^+^ cells, suggesting the absence of innate lymphoid cells from control or injured human nerve tissues.^33^

Lastly, we found the area fractions of intraneural MBP, but not β-tubulin III, significantly correlated with the densities of intraneural CD3^+^ and CD68^+^ cells in Morton’s neuroma (Supplemental Table 9, Supplemental Figure 3).

## Discussion

In this study evaluating gene expression profiles in human injured and control nerves, a large number of genes were found to be differentially expressed. Both GO and WGCNA analyses identified biological processes involved with a response of the immune system and inflammation and development of the nervous system (e.g., myelination, axon guidance) to be overrepresented. Deconvolution analysis confirmed differences in immune cell populations between Morton’s neuroma and control nerves with macrophage proportions not only greater in Morton’s neuroma but also correlating with the severity of paroxysmal pain. Of note, we identified an overexpressed gene signature (*MARCO, CD163* and *STAB1*) associated with a specific subset of M(GC) macrophages, with the expression of *MARCO* being correlated with the severity of paroxysmal pain. Immunohistochemical analysis in nerve sections validated the presence of demyelination, intraneural increase in T-cells and M(GC) MARCO^+^ subset macrophages in Morton’s neuroma samples, while CD68^+^ macrophage density correlated with burning pain. These findings implicate an ongoing role for the immune system in the context of chronic peripheral neuropathic pain in humans, with a potentially interesting role for macrophages and specifically the MARCO^+^ M(GC) subset.

Our findings clearly highlight a role of local neuroinflammation in the context of chronic neuropathic pain in humans. Neuroinflammation and its importance in the initiation and maintenance of neuropathic pain is well established in preclinical models,^34,35^ with a particular prominence of macrophages and T-cells^36,37^ but also neutrophils and mast cells. However, evidence for its role in human nerve injuries has remained largely circumstantial due to challenges of access to tissues.^13^ Most evidence to date stems from indirect methods such as systemic blood/cerebrospinal fluid analysis^38,39^ with histological analyses of nerve biopsies only performed in very severe and debilitating conditions that justify nerve biopsy (e.g. vasculitic neuropathies or chronic axonal neuropathies).^40,41^ Whereas these studies support a role of inflammation in severe neuropathic pain conditions in humans, our current findings suggest that milder forms of nerve injuries such as entrapment neuropathies are also associated with neuroinflammation. This is in line with our previous preclinical findings of local immune cell activation in a model of mild chronic nerve compression.^42^

Of importance, the patient population studied here had longstanding symptoms associated with nerve injury (median duration 2.5 years). Most pre-clinical models study the effect of neuroinflammation over relatively short periods in the order of weeks rather than years after onset of nerve injury.^43^ These studies suggest that local densities of immune cells remain high for up to 14 weeks after nerve ligation,^37^ 7 weeks after chronic constriction injury^44^ and 12 weeks after mild nerve compression.^42^ However, the scarcity of long-term studies precludes in-depth knowledge about the exact timing of neuroinflammation persistence and its resolution.

Our data highlight that neuroinflammation continues to play a role in chronic neuropathic pain, long after the initial onset of nerve injury. This could be either attributed to continuing axon or myelin degeneration due to ongoing nerve compression, or the failure of inflammation to resolve. Indeed, even though *CD163* (expressed on anti-inflammatory macrophages) was high at molecular and cellular level, pro-resolution markers such as *Arginase1* or *FoxP3*, were either absent or very lowly expressed in our dataset, replicating what has been reported in mouse.^37^ The importance of resolution of neuroinflammation has gained increasing interest in the past decade with studies highlighting insufficiency or dysregulation of pain-resolving immune cells and their mediators in experimental neuropathic pain.^36^ There is also emerging evidence for the pro-resolution machinery as an important player in human chronic pain persistence^45^ and resolution.^46^ A dysfunction of resolution may explain why steroid injections are only short-lasting in patients with Morton’s neuroma,^47^ and other entrapment neuropathies.^48^

Screening of differentially expressed genes associated with co-expression modules identified consistent upregulation of three genes (*MARCO, STAB1* and *CD163*) associated with a specific subset of M(GC) glucocorticoid-induced macrophages.^32^ MARCO (= macrophage receptor with collagenous structure) is a class A scavenger receptor, which has been implicated in opsonin-independent phagocytosis, cell signalling in inflammation, and macrophages that adopt a pro-fibrotic phenotype.^49–51^ MARCO expression can be upregulated within hypoxic microenvironments,^52^ such as found in peripheral neuropathies.^53,54^ We identified a correlation between *MARCO* expression and paroxysmal pain, although this was not replicated with immunohistological staining. It could be speculated that the stark upregulation of this type of macrophage reflects a continuing but potentially failing effort to combat the ongoing nerve compression and hypoxic intraneural environment. Alternatively, it could be that the presence of MARCO+ M(GC) macrophages indicates cells which are promoting fibrosis in a non-resolution immune environment^49^ – thus preventing a return to pain-free tissue homeostasis. Finally, our identification of this subset raises the question whether steroid injections might not only be ineffective as a treatment long-term, but perhaps might even promote the persistence of this potentially counter-productive macrophage population.

In addition to a strong inflammation signature, our data also implicates processes related to the development of the nervous system. It is well established that nerve injuries are associated with demyelination, which persists to chronic stages of human entrapment neuropathies.^55,56^ The immunohistochemical experiments validated the myelination gene signature, with a stark reduction in myelin basic protein staining in Morton’s neuromas.

In Morton’s neuroma sections, we also observed intraneural areas which lacked axons, presumably indicating fibrotic changes (see also upregulated GO terms collagen fibril organisation and cellular response to transforming growth factor beta stimulus). Fibrotic contribution is well established in other entrapment neuropathies such as carpal tunnel syndrome, where fibrotic factors such as TGF-beta are strongly upregulated.^57–59^ However, early non-quantitative studies suggest that similar fibrotic areas are present in non-painful plantar nerves.^60,61^ It could be the case that fibrosis is only pro-algesic if it is accompanied by a pro-inflammatory environment, as is the case in our Morton’s samples.

Finally, while quantification of overall anti-β-tubulin III staining was not altered in Morton’s neuroma compared to control nerves, it negatively correlated with pain, corroborating the findings from the WGCNA analysis where the neurogenesis module negatively correlated with paroxysmal pain.

## Limitations

Several limitations should be considered when interpreting our findings. First, location of control human nerves varied, with only three samples taken at the same anatomical site as the Morton’s neuroma due to a lack of tissue availability. We deemed it more important to collect fresh human nerve rather than post-mortem nerves due to RNA degradation and age influences on peripheral nerve integrity and inflammatory processes. Differences in nerve composition (e.g., motor vs sensory) may have influenced our findings. Based on the PCA plots, the different types of nerves did not cluster together, making this unlikely. The three control nerves collected from amputees are more similar to Morton’s neuroma than the other control samples. This means that including both amputees and non-amputees might have reduced the effect sizes in Morton’s neuroma vs control nerves. Second, whereas our age matching was successful, the proportions of male and female participants were disparate in the two groups, reflecting the clinical populations of Morton’s neuroma (female dominant)^62^ versus flap or amputation surgeries often secondary to trauma (male dominant). The relatively small sample size prohibited separate analyses of male and female nerve samples. Sex differences related to immune function^63^ and pain^64^ are well established and recent evidence from healthy human peripheral nerves also reported sexually dimorphic gene expressions.^14^ In this study we considered and controlled for an additive effect of gender in Morton’s neuroma vs control nerves. Gender is not one of the primary outcomes, we rather blocked for its effect on Morton’s neuroma vs control nerves, so we do not report results for this coefficient. Third, the use of human tissue comes with inherent variation (e.g., chronicity, severity). While we were careful to exclude potential co-morbidities that may influence nerve health in both groups, human tissue variation is likely to mask more subtle effects. Of note, this increases confidence in the identified changes, which are likely of considerable size to survive background noise. Lastly, we only had a limited amount of human tissue, and concentrated on validating the key findings from our bulk sequencing experiment.

## Conclusions

In conclusion, our combined transcriptomic and histological findings suggest an intraneural signature related to inflammation and development of the nervous system (e.g., myelination, neurogenesis) to be associated with Morton’s neuroma. We identified correlations between deconvolution-derived immune cell populations, WGCNA modules and single genes (*MARCO*) with paroxysmal pain, while histological CD68+ macrophage density correlated with burning pain. These findings implicate neuroinflammation in the chronic stage of human neuropathic pain associated with focal nerve injury. The M(GC) MARCO^+^ subset of macrophages may be of interest in the context of paroxysmal pain.

## Acknowledgement

We would like to thank all participants for taking part in this study. The help of the Clinical Research Network Nurses at Oxford University Hospitals NHS Foundation Trust and Royal Berkshire NHS Foundation Trust as well as the biobanking team at Balgrist Campus is gratefully acknowledged. We also thank all surgeons for helping with intraoperative sample collection. We thank the High-Throughput Genomics Group at the Wellcome Trust Centre for Human Genetics (funded by Wellcome Trust grant reference 090532/Z/09/Z) for the generation of the sequencing data.

## Funding information

ABS is supported by a Wellcome Trust Clinical Career Development Fellowship (222101/Z/20/Z). IDS and PSS were funded by the Clinical Research Priority Program (CRPP) Pain from the University of Zurich, Switzerland. PSS is supported by postdoctoral fellowships from the International Foundation for Research in Paraplegia (P198F), the Swiss National Science Foundation (P500PB_214416), and Michael Smith Health Research BC (RT-2023-3173). This research was funded in whole, or in part, by the Wellcome Trust [222101/Z/20/Z]. For the purpose of Open Access, the author has applied a CC BY public copyright license to any Author Accepted Manuscript version arising from this submission. We also received funding from ONO pharamceuticals Ltd.

## Competing interests

The authors report no conflict of interest. The funders had no input to the design, analysis or interpretation of the study.

## Supplementary material

Supplementary material is available at *Brain* online

## Notes

### Competing Interest Statement

The authors have declared no competing interest.

### Author Declarations

Ethics Committee South Central - Oxford C Research Ethics Committee gave ethical approval for this research (REF 18/SC/0410). Ethics Committee London - Camden & Kings Cross Research Ethics Committee gave ethical approval for this research (REC16/LO/1920) Ethics committee of Canton of Zurich gave ethical approval for this work (2018-02198).

