## Supplemental Files and Figures for "The Molecular Signature of Neuropathic Pain in a Human Model System"

**Supplemental Table 1: Primary and secondary antibodies used in the study**

| **Primary antibody to** | **Host** | **Clonality** | **Isotype** | **Conjugated** | **Dilution** | **Catalogue no.** | **Company** |
| --- | --- | --- | --- | --- | --- | --- | --- |
| Human myelin basic protein | Rat | Monoclonal | IgG2a | N/A | 1:500 | ab7349 | Abcam |
| Human β-tubulin III | Rabbit | Polyclonal | IgG | N/A | 1:500 | T2200 | Sigma-Aldrich |
| Human CD3 | Rabbit | Monoclonal | IgG | N/A | 1:100 | ab16669 | Abcam |
| Human CD68 | Mouse | Monoclonal | IgG1 | N/A | 1:200 | ab955 | Abcam |
| Human CD3 | Mouse | Monoclonal | IgG2a | N/A | 1:25 | ab699 | Abcam |
| Human CD4 | Rabbit | Monoclonal | IgG | N/A | 1:50 | ab133616 | Abcam |
| Human CD163 | Mouse | Monoclonal | IgG1 | N/A | 1:100 | NB-40686AF647 | Bio-techne |
| Human MARCO | Rabbit | Polyclonal | IgG | N/A | 1:50 | PA5-64134 | Invitrogen |
| **Secondary antibody to** | **Host** | **Clonality** | **Isotype** | **Conjugated** | **Dilution** | **Catalogue no.** | **Company** |
| Rabbit IgG (H+L) | Donkey | Polyclonal | IgG | AF546 | 1:500 | A10040 | Invitrogen |
| Rat IgG (H+L) | Goat | Polyclonal | IgG | AF488 | 1:500 | A11006 | Invitrogen |
| Mouse IgG (H+L) | Goat | Polyclonal | IgG | AF546 | 1:500 | A11003 | Invitrogen |
| Rabbit IgG (H+L) | Goat | Polyclonal | IgG | AF488 | 1:500 | A11008 | Invitrogen |

**Supplemental Table 2: List of genes and their relative expression values denoted as ‘M1’ and ‘M2’ macrophages in the deconvolution analysis**

| LM22 gene signatures | | |
| --- | --- | --- |
| GENE SYMBOL | LM22 Macrophages_M1 | LM22 Macrophages_M2 |
| *ABCB4* | 27.55771 | 121.432277 |
| *ACAP1* | 30.974254 | 23.05515 |
| *ACHE* | 1373.06962 | 48.585761 |
| *ACP5* | 1062.475909 | 7633.960182 |
| *AIF1* | 1611.228571 | 11610.29035 |
| *AIM2* | 2965.888162 | 148.284522 |
| *ALOX15* | 5.412216 | 430.335305 |
| *ALOX5* | 83.300898 | 248.372472 |
| *ANGPT4* | 21.218127 | 11.809701 |
| *APOBEC3A* | 4501.140479 | 148.79934 |
| *APOL3* | 11009.29392 | 437.527208 |
| *APOL6* | 967.645146 | 7.784612 |
| *AQP9* | 4309.341662 | 399.474619 |
| *ASGR1* | 25.236261 | 155.491339 |
| *ASGR2* | 26.373869 | 760.725771 |
| *ATP8B4* | 33.351428 | 1163.254876 |
| *BANK1* | 16.020271 | 29.192635 |
| *BHLHE41* | 143.868108 | 325.12977 |
| *BIRC3* | 1971.112539 | 237.501784 |
| *BPI* | 16.533686 | 25.96026 |
| *BST1* | 73.768331 | 209.464822 |
| *C5AR1* | 262.803764 | 1920.668117 |
| *CCDC102B* | 1.538368 | 2.55642 |
| *CCL13* | 182.28391 | 6581.432651 |
| *CCL14* | 1233.172359 | 2459.096418 |
| *CCL17* | 55.411581 | 620.213461 |
| *CCL18* | 1252.527933 | 12334.37123 |
| *CCL19* | 32553.88074 | 137.074204 |
| *CCL20* | 724.614868 | 12.804997 |
| *CCL22* | 464.972961 | 880.202204 |
| *CCL23* | 615.574234 | 5505.223986 |
| *CCL5* | 16862.16904 | 250.750217 |
| *CCL7* | 241.065018 | 85.74723 |
| *CCL8* | 4535.517152 | 2829.09296 |
| *CCND2* | 81.430532 | 108.189616 |
| *CCR2* | 80.9645 | 74.521205 |
| *CCR5* | 1493.347557 | 787.300008 |
| *CCR7* | 17366.91051 | 123.746312 |
| *CD180* | 120.080425 | 2787.643912 |
| *CD1C* | 22.859693 | 167.50326 |
| *CD1D* | 373.059453 | 39.50876 |
| *CD209* | 190.945084 | 2758.297802 |
| *CD244* | 46.758659 | 96.281434 |
| *CD300A* | 211.104829 | 1737.241257 |
| *CD37* | 340.016978 | 2337.797784 |
| *CD38* | 12009.36242 | 642.610487 |
| *CD4* | 1574.336844 | 10105.34674 |
| *CD40* | 12966.60573 | 643.639203 |
| *CD7* | 13.368126 | 80.617694 |
| *CD72* | 9.588069 | 123.95208 |
| *CD80* | 3650.811207 | 160.481079 |
| *CDA* | 8.817497 | 28.81247 |
| *CFP* | 219.1912 | 1971.112539 |
| *CHI3L1* | 3395.663462 | 2050.816822 |
| *CHI3L2* | 1765.552386 | 175.383593 |
| *CHST15* | 103.487155 | 1051.6856 |
| *CLCA3P* | 0.251594 | 9.029404 |
| *CLEC10A* | 69.396554 | 5770.127086 |
| *CLEC2D* | 1465.741277 | 187.986571 |
| *CLEC4A* | 51.678716 | 6742.94324 |
| *CLEC7A* | 283.203808 | 3071.447657 |
| *CLIC2* | 1393.382865 | 2233.164343 |
| *COL8A2* | 33.843537 | 151.562355 |
| *CREB5* | 11.762749 | 59.217662 |
| *CRTAM* | 38.222697 | 85.708983 |
| *CRYBB1* | 12.127181 | 166.955398 |
| *CSF1* | 35.288498 | 23.122173 |
| *CSF3R* | 860.337222 | 737.859904 |
| *CXCL10* | 31273.67963 | 278.343404 |
| *CXCL11* | 9059.841342 | 29.35941 |
| *CXCL13* | 627.930517 | 16.760769 |
| *CXCL3* | 271.246673 | 233.330355 |
| *CXCL5* | 91.980538 | 46.38135 |
| *CXCL9* | 38944.01868 | 199.435917 |
| *CXCR5* | 212.275007 | 14.431145 |
| *CYP27A1* | 72.621008 | 75.710854 |
| *CYP27B1* | 6662.564771 | 145.701121 |
| *DACH1* | 4.273622 | 8.232486 |
| *DCSTAMP* | 171.556253 | 278.806442 |
| *DHRS11* | 18.600266 | 242.690661 |
| *DHX58* | 1394.307066 | 233.583127 |
| *DPEP2* | 16.216382 | 2410.189476 |
| *EBI3* | 20437.69231 | 311.205868 |
| *EGR2* | 23.087546 | 2175.756551 |
| *ELANE* | 103.353302 | 206.315658 |
| *EPB41* | 7.165488 | 25.556869 |
| *FCER1A* | 56.548697 | 801.886611 |
| *FCER2* | 10.526179 | 352.928032 |
| *FCN1* | 306.829655 | 593.095894 |
| *FES* | 20.942158 | 982.200085 |
| *FOSB* | 23.170426 | 22.497432 |
| *FPR1* | 801.886611 | 408.541527 |
| *FPR2* | 274.936574 | 95.438646 |
| *FPR3* | 608.269253 | 2357.542622 |
| *FRK* | 3.762991 | 3.129846 |
| *FRMD4A* | 256.173231 | 1094.097795 |
| *FZD3* | 32.632427 | 195.35077 |
| *GGT5* | 290.215074 | 407.794036 |
| *GPC4* | 129.318831 | 145.871911 |
| *GPR1* | 0.972465 | 0.453354 |
| *GPR183* | 1631.804333 | 2474.434424 |
| *GUSBP11* | 14.61436 | 286.719961 |
| *HAL* | 90.959858 | 76.240189 |
| *HESX1* | 2324.301051 | 100.313358 |
| *HHEX* | 66.505428 | 566.324939 |
| *HK3* | 31.9761 | 159.121705 |
| *HLA-DOB* | 1573.187324 | 41.576516 |
| *HLA-DQA1* | 4169.716387 | 5225.867112 |
| *HPGDS* | 64.876999 | 138.674103 |
| *HRH1* | 145.283481 | 2195.073255 |
| *HSPA6* | 139.862714 | 207.498138 |
| *HTR2B* | 31.869533 | 216.050282 |
| *IDO1* | 26473.42255 | 91.367926 |
| *IFI44L* | 6016.733404 | 984.730251 |
| *IGHM* | 32.677241 | 26.518324 |
| *IGSF6* | 295.072409 | 1626.552661 |
| *IL12B* | 513.084829 | 18.653005 |
| *IL18RAP* | 24.975232 | 38.640956 |
| *IL1A* | 90.023426 | 27.047524 |
| *IL1B* | 675.121823 | 291.824541 |
| *IL2RA* | 3963.774053 | 146.46007 |
| *IL7R* | 2163.365124 | 85.591441 |
| *LAG3* | 1427.702674 | 97.677236 |
| *LAMP3* | 17116.19562 | 113.141386 |
| *LAT* | 162.813157 | 181.952672 |
| *LILRA2* | 317.685623 | 220.178832 |
| *LTA* | 114.104195 | 14.981975 |
| *LTC4S* | 145.924869 | 581.668952 |
| *LY9* | 28.294121 | 252.279981 |
| *MAK* | 40.723703 | 106.229954 |
| *MAP4K1* | 31.103853 | 367.271055 |
| *MARCO* | 48.772663 | 117.151134 |
| *MEFV* | 10.082345 | 9.173433 |
| *MEP1A* | 25.482354 | 66.466162 |
| *MGAM* | 5.166818 | 5.665802 |
| *MMP9* | 2727.981464 | 16386.05747 |
| *MNDA* | 4016.523813 | 1867.617958 |
| *MS4A6A* | 402.562324 | 16167.182 |
| *MSC* | 1562.138826 | 101.484341 |
| *NCF2* | 762.035112 | 11487.88902 |
| *NFE2* | 32.428804 | 110.075989 |
| *NKG7* | 217.224577 | 6.547728 |
| *NLRP3* | 37.679105 | 322.062046 |
| *NME8* | 97.090451 | 422.959594 |
| *NPL* | 501.961637 | 4287.536344 |
| *P2RX5* | 21.687396 | 20.011502 |
| *P2RY13* | 473.333504 | 2965.888162 |
| *P2RY14* | 21.358531 | 852.556374 |
| *P2RY2* | 11.441462 | 9.46357 |
| *PADI4* | 17.759004 | 12.531005 |
| *PAQR5* | 18.280857 | 17.469058 |
| *PCDHA5* | 85.938604 | 8.255798 |
| *PIK3IP1* | 209.464822 | 1631.804333 |
| *PLA1A* | 2799.769624 | 14.403984 |
| *PLA2G7* | 1679.064856 | 1569.463396 |
| *PLEKHF1* | 73.439523 | 33.492488 |
| *PPBP* | 230.145918 | 292.991822 |
| *PPFIBP1* | 6.90617 | 11.475809 |
| *QPCT* | 113.835212 | 1278.360026 |
| *RASGRP2* | 120.640212 | 102.042176 |
| *RASGRP3* | 552.034418 | 1320.702754 |
| *REPS2* | 9.789026 | 171.230898 |
| *RNASE2* | 407.22257 | 677.713898 |
| *RRP12* | 11.728149 | 14.514662 |
| *RSAD2* | 6976.481671 | 140.167353 |
| *S100A12* | 258.300428 | 123.701236 |
| *SEC31B* | 58.50851 | 492.069202 |
| *SELL* | 234.171636 | 287.219139 |
| *SIGLEC1* | 6220.794765 | 3457.591061 |
| *SLAMF1* | 6057.915205 | 845.506509 |
| *SLAMF8* | 663.473323 | 1407.517431 |
| *SLC12A8* | 24.556462 | 25.339354 |
| *SLC2A6* | 6057.915205 | 579.733612 |
| *SLCO5A1* | 954.038207 | 39.757128 |
| *SOCS1* | 2300.259226 | 705.135411 |
| *SPIB* | 1435.057428 | 31.210885 |
| *ST3GAL6* | 41.075062 | 192.217977 |
| *ST6GALNAC4* | 133.983253 | 243.299101 |
| *STAP1* | 372.920945 | 43.53483 |
| *STEAP4* | 22.350852 | 26.463843 |
| *TARDBPP1* | 4.672469 | 6.606825 |
| *TCF7* | 284.03179 | 43.911528 |
| *TEP1* | 69.867938 | 112.195761 |
| *TMEM255A* | 289.163953 | 167.015226 |
| *TNFAIP6* | 19190.61594 | 325.364338 |
| *TNFRSF10C* | 42.180709 | 75.929976 |
| *TNFRSF11A* | 74.820238 | 396.39139 |
| *TNFRSF4* | 1462.50333 | 441.126154 |
| *TNFSF14* | 20.454372 | 16.045444 |
| *TNIP3* | 1192.706587 | 17.3641 |
| *TREM1* | 194.725477 | 184.649791 |
| *TREM2* | 160.831659 | 3999.017687 |
| *TRIB2* | 71.304648 | 22.363741 |
| *UPK3A* | 6.818438 | 7.606821 |
| *VNN1* | 12.28114 | 120.640212 |
| *VNN2* | 6.198523 | 8.255798 |
| *WNT5B* | 11.169494 | 533.28442 |
| *ZBTB32* | 223.024334 | 14.981975 |
| *ZFP36L2* | 23.572098 | 303.576954 |
| *ZNF135* | 4.993364 | 39.757128 |

| IMMST gene signatures | | |
| --- | --- | --- |
| GENE Symbol | IMMST macrophage_m1 | IMMST macrophage_m2 |
| *ABCA5* | 24.630897 | 24.961502 |
| *ABCB1* | 15.464132 | 13.566988 |
| *ABHD5* | 108.259229 | 85.51079 |
| *ADAM19* | 36.512586 | 33.11254 |
| *ADAMTS5* | 10.081041 | 9.472768 |
| *ADI1* | 48.616494 | 79.643998 |
| *AGPAT5* | 50.099996 | 42.497776 |
| *ALAS1* | 87.283585 | 64.273069 |
| *ANXA3* | 11.542702 | 8.375526 |
| *APOBEC3A* | 263.131611 | 37.612708 |
| *APOBEC3G* | 114.312067 | 47.078507 |
| *APOE* | 770.980636 | 468.985784 |
| *ARID4A* | 62.114806 | 77.447609 |
| *ARNT2* | 16.374199 | 9.575317 |
| *ASGR1* | 81.902272 | 96.547202 |
| *ASGR2* | 30.691987 | 66.222538 |
| *ASRGL1* | 31.970687 | 45.722372 |
| *ATP2B1* | 40.32152 | 63.084047 |
| *C1QA* | 183.454527 | 174.396667 |
| *C1QB* | 760.979895 | 415.591757 |
| *CACNA2D3* | 14.996063 | 16.45717 |
| *CALML4* | 52.953172 | 61.126032 |
| *CAMK4* | 10.568397 | 9.461182 |
| *CASP1* | 227.559694 | 128.462902 |
| *CCR2* | 19.011265 | 26.252999 |
| *CCR3* | 13.944134 | 15.410815 |
| *CD14* | 1312.160328 | 544.111938 |
| *CD163* | 323.425414 | 326.252005 |
| *CD19* | 30.032093 | 30.371927 |
| *CD1D* | 34.051541 | 24.184756 |
| *CD1E* | 22.479124 | 79.399689 |
| *CD207* | 11.735855 | 15.369275 |
| *CD209* | 180.567087 | 642.842597 |
| *CD38* | 341.629037 | 31.603112 |
| *CD3D* | 19.449839 | 16.652012 |
| *CD4* | 105.571924 | 152.733516 |
| *CD8A* | 18.086649 | 18.981875 |
| *CDC14B* | 19.040319 | 14.904207 |
| *CDC42EP4* | 118.310048 | 86.283577 |
| *CDK2AP2* | 50.337277 | 53.60228 |
| *CDKN1C* | 23.53832 | 22.066926 |
| *CDR2L* | 23.844203 | 76.287923 |
| *CEACAM3* | 23.547452 | 22.994622 |
| *CES1* | 83.84816 | 43.647947 |
| *CFB* | 468.539663 | 64.509487 |
| *CHMP7* | 45.414453 | 42.674649 |
| *CHST7* | 129.464542 | 145.295908 |
| *CIB2* | 24.881341 | 20.905479 |
| *CLCF1* | 74.896664 | 46.968353 |
| *CLEC4E* | 58.537679 | 20.167679 |
| *CLEC5A* | 109.754963 | 39.816376 |
| *CLIC2* | 73.083241 | 124.119722 |
| *CNNM1* | 28.06154 | 29.605258 |
| *CNOT1* | 82.160237 | 94.498375 |
| *CP* | 26.831256 | 13.188198 |
| *CRISPLD2* | 78.675672 | 22.916827 |
| *CRLF2* | 17.549464 | 15.361947 |
| *CSTA* | 97.222436 | 85.573577 |
| *CXCL10* | 950.98551 | 24.653657 |
| *CXCL9* | 557.824855 | 26.517111 |
| *CYCS* | 56.353643 | 70.424246 |
| *CYP4F3* | 14.912731 | 10.663558 |
| *CYSLTR1* | 47.664642 | 52.269631 |
| *DEFB1* | 51.553472 | 16.54388 |
| *DENND3* | 75.596479 | 51.239597 |
| *DPYD* | 136.567867 | 160.421642 |
| *DUSP4* | 15.619161 | 15.879018 |
| *DYSF* | 81.318775 | 62.847265 |
| *EDN1* | 222.128369 | 41.451488 |
| *EIF1* | 1127.387628 | 1147.809467 |
| *ELSPBP1* | 12.417795 | 9.097566 |
| *EMILIN2* | 559.149007 | 596.840208 |
| *EVL* | 325.23282 | 792.525801 |
| *FCHO1* | 52.11439 | 67.194768 |
| *FCN1* | 507.278422 | 357.450773 |
| *FGFR3* | 21.19645 | 20.901608 |
| *FGR* | 230.896904 | 224.091163 |
| *FLT4* | 9.057131 | 10.352679 |
| *FMO5* | 25.903274 | 24.73977 |
| *FST* | 38.960743 | 35.174874 |
| *FSTL1* | 16.63998 | 14.025324 |
| *FUT3* | 14.130251 | 15.878089 |
| *FXYD6* | 71.340161 | 36.164957 |
| *GAS7* | 43.743887 | 56.949024 |
| *GATA2* | 13.724963 | 11.863059 |
| *GBP1* | 896.719452 | 71.273239 |
| *GCH1* | 137.261667 | 19.51714 |
| *GFOD1* | 85.044203 | 121.400191 |
| *GIMAP4* | 396.81946 | 166.154557 |
| *GLRA1* | 10.249014 | 10.443668 |
| *GPNMB* | 1031.714985 | 1121.365578 |
| *HAGH* | 157.716157 | 156.487197 |
| *HAVCR1* | 7.744264 | 7.164532 |
| *HBD* | 12.990704 | 12.819894 |
| *HCK* | 534.413194 | 408.408765 |
| *HDC* | 10.868859 | 12.853772 |
| *HOMER2* | 17.154798 | 124.184141 |
| *HPSE* | 52.394734 | 33.544332 |
| *HTRA1* | 42.297892 | 38.709417 |
| *IFI27* | 605.424453 | 34.217672 |
| *IFNB1* | 11.83225 | 8.995349 |
| *IFT20* | 42.159323 | 65.158177 |
| *IL15RA* | 66.526337 | 17.317901 |
| *IL1R2* | 31.41673 | 54.540967 |
| *IL2RA* | 68.765118 | 21.081191 |
| *IL32* | 69.334626 | 21.187101 |
| *IL5RA* | 23.854369 | 22.964176 |
| *IL6ST* | 36.237525 | 34.355542 |
| *ING2* | 108.432819 | 121.012868 |
| *IRS1* | 13.367414 | 12.076048 |
| *JRKL* | 29.091427 | 28.107682 |
| *KCNJ15* | 19.471191 | 13.970518 |
| *KIF1A* | 10.596884 | 9.506352 |
| *KIF22* | 22.122568 | 20.208357 |
| *KLHL18* | 52.583207 | 51.647409 |
| *KLRC3* | 10.861977 | 9.579869 |
| *KLRF1* | 21.005789 | 25.999136 |
| *KRT5* | 10.378597 | 10.101124 |
| *KSR1* | 23.544305 | 18.866856 |
| *LAIR1* | 413.762792 | 302.280213 |
| *LAMP3* | 133.221992 | 47.03768 |
| *LILRA4* | 33.753721 | 41.414371 |
| *LILRA5* | 111.697649 | 44.101029 |
| *LILRB1* | 283.362864 | 206.378371 |
| *LIMA1* | 74.208216 | 149.422444 |
| *LIMK2* | 54.284855 | 21.660486 |
| *LIPF* | 23.402844 | 20.990309 |
| *LRP5L* | 61.170911 | 74.419544 |
| *LRRC8D* | 81.105576 | 89.427665 |
| *LSM4* | 192.355209 | 205.887501 |
| *MAG* | 14.515489 | 14.017755 |
| *MAL* | 16.023397 | 24.780659 |
| *MAOA* | 70.411593 | 476.221065 |
| *MAPK7* | 51.413285 | 48.983391 |
| *MAT2B* | 130.12854 | 175.130871 |
| *MEST* | 11.403056 | 11.188716 |
| *MMP25* | 49.681395 | 41.162143 |
| *MMP8* | 7.394264 | 6.948941 |
| *MMP9* | 3825.404778 | 2402.56263 |
| *MOCS3* | 25.069081 | 27.340857 |
| *MPO* | 45.855922 | 42.880359 |
| *MPPED2* | 6.65776 | 6.769345 |
| *MRPL3* | 111.359692 | 165.734104 |
| *MRPL4* | 43.657143 | 45.236235 |
| *MS4A1* | 12.435397 | 13.626376 |
| *MS4A6A* | 84.453534 | 675.750915 |
| *MT1X* | 624.849502 | 146.602072 |
| *MTMR11* | 45.061238 | 33.356051 |
| *MTSS1* | 53.515305 | 86.251774 |
| *MUC1* | 61.029106 | 27.572684 |
| *MYLIP* | 105.479133 | 111.74344 |
| *NAGA* | 82.301948 | 150.593305 |
| *NBN* | 107.383916 | 69.558193 |
| *NBR1* | 20.301084 | 32.997768 |
| *NDRG2* | 48.461579 | 52.430672 |
| *NOTCH4* | 25.50794 | 16.457332 |
| *NPEPPS* | 59.871167 | 74.879931 |
| *NR2E3* | 10.350706 | 10.206289 |
| *NR4A2* | 49.373216 | 34.249654 |
| *NRG1* | 11.224971 | 11.929372 |
| *NRGN* | 24.088339 | 47.573654 |
| *NUDT1* | 40.586782 | 64.728852 |
| *NUDT18* | 105.166233 | 81.773497 |
| *NXT1* | 187.901648 | 191.35917 |
| *NXT2* | 94.351477 | 85.265605 |
| *OLFM1* | 23.836636 | 21.128798 |
| *OR2F1* | 9.727019 | 9.947481 |
| *OR2J2* | 8.206225 | 8.262144 |
| *ORM1* | 71.607499 | 15.634066 |
| *OSBPL10* | 16.201821 | 23.093357 |
| *PALLD* | 64.220082 | 322.839532 |
| *PANX1* | 93.926979 | 38.536156 |
| *PAX5* | 12.432153 | 10.260643 |
| *PCGF2* | 54.521158 | 38.416585 |
| *PDGFB* | 31.85218 | 39.577951 |
| *PDK4* | 32.039133 | 41.872915 |
| *PGLYRP1* | 16.973305 | 17.564939 |
| *PI3* | 18.590268 | 12.491856 |
| *PIK3CG* | 85.420994 | 81.712739 |
| *PLAT* | 14.389554 | 11.157077 |
| *POU2AF1* | 13.441854 | 16.72148 |
| *PPA1* | 883.864571 | 284.584418 |
| *PROM1* | 8.904687 | 12.974965 |
| *PSAT1* | 27.617124 | 24.651705 |
| *PTPN13* | 10.596448 | 11.97721 |
| *PTTG2* | 47.705921 | 43.386717 |
| *QPRT* | 47.82859 | 262.219569 |
| *RAB9A* | 334.077305 | 314.48347 |
| *RAMP1* | 22.138471 | 192.592609 |
| *RETN* | 15.702533 | 13.807824 |
| *RNASE1* | 98.189993 | 293.296595 |
| *RNASE4* | 40.494257 | 42.010005 |
| *RNF122* | 33.337548 | 31.926228 |
| *RPL7* | 121.95721 | 185.192881 |
| *RRAS* | 271.274825 | 203.627594 |
| *S100A12* | 31.095649 | 13.012204 |
| *S100B* | 10.358643 | 11.899717 |
| *S100P* | 27.529592 | 20.798301 |
| *SCRN1* | 73.293637 | 90.385334 |
| *SERPINF2* | 22.968984 | 27.100463 |
| *SETBP1* | 17.205978 | 20.812313 |
| *SF3A3* | 146.546483 | 184.997574 |
| *SFTPD* | 11.760854 | 17.679207 |
| *SFXN3* | 49.675081 | 52.549458 |
| *SH3BP2* | 29.562566 | 28.184269 |
| *SIDT1* | 14.048635 | 13.890627 |
| *SIGLEC6* | 17.207657 | 20.742556 |
| *SLC12A3* | 11.621312 | 11.100721 |
| *SLC15A3* | 1508.414675 | 1224.507916 |
| *SLC17A5* | 23.011273 | 37.038666 |
| *SLC1A4* | 117.395652 | 53.088537 |
| *SLC4A1AP* | 73.529014 | 71.101557 |
| *SLC6A13* | 41.57943 | 36.114688 |
| *SLC7A7* | 1257.300977 | 647.250127 |
| *SLC9A3R1* | 115.444262 | 97.944347 |
| *SLCO2B1* | 144.379303 | 353.058568 |
| *SMARCD3* | 38.19507 | 36.757718 |
| *SOCS2* | 51.89447 | 30.662962 |
| *STAB2* | 9.210147 | 7.147536 |
| *STEAP4* | 12.781317 | 13.029812 |
| *SYNE1* | 15.641658 | 19.558229 |
| *TAGLN* | 47.473968 | 43.976212 |
| *TBC1D8* | 89.97544 | 75.585088 |
| *TCL1A* | 14.626984 | 13.342963 |
| *TFEC* | 44.374457 | 53.039233 |
| *TLL1* | 7.336204 | 7.513473 |
| *TLR5* | 42.20772 | 96.719577 |
| *TLR8* | 160.354789 | 88.426538 |
| *TM4SF5* | 7.056287 | 7.145807 |
| *TMC6* | 141.064862 | 189.397456 |
| *TNFRSF13B* | 16.406338 | 15.627486 |
| *TNFRSF25* | 13.7575 | 18.546814 |
| *TNNI2* | 36.851402 | 42.340209 |
| *TOMM22* | 126.506229 | 134.698281 |
| *TRAF3IP2* | 53.077523 | 29.147313 |
| *TRAT1* | 13.846022 | 10.402862 |
| *TREM1* | 60.239799 | 36.983424 |
| *TRIB1* | 351.065142 | 440.992267 |
| *TSPAN7* | 10.389817 | 42.686842 |
| *TUBB6* | 174.303105 | 203.20431 |
| *TULP2* | 8.486431 | 10.022635 |
| *UBE2J1* | 225.380084 | 187.877026 |
| *ULK2* | 43.445832 | 37.428174 |
| *WEE1* | 29.493918 | 22.905962 |
| *ZC3H12A* | 324.661191 | 132.978126 |
| *ZDHHC13* | 44.326729 | 37.377875 |
| *ZNF180* | 11.159899 | 10.36369 |
| *ZNF189* | 51.875213 | 47.874599 |
| *ZNF34* | 42.127202 | 48.465928 |
| *ZNF593* | 113.078329 | 95.599543 |

**Supplemental Table 3: Correlations of top enriched GO terms and selected gene markers of M(GC) MARCO^+^ macrophages with clinical pain phenotype**

| **Top enriched GO term** | **NPSI burning** | **NPSI deep** | **NPSI paroxysmal** | **NPSI evoked** | **NPSI paraesthesia** | **NPSI total** | **VAS pain 24H** | **VAS numbness 24H** |
| --- | --- | --- | --- | --- | --- | --- | --- | --- |
| immune response | 0.07 | 0.28 | 0.31 | 0.32 | -0.01 | 0.21 | 0.03 | -0.06 |
| collagen metabolic process | 0.15 | 0.14 | 0.37 | 0.1 | 0.01 | 0.15 | 0.09 | -0.12 |
| defense response | 0.02 | 0.11 | **0.66*** | 0.04 | 0.24 | 0.25 | 0.09 | -0.16 |
| muscle system process | 0.18 | -0.14 | -0.29 | **-0.44*** | -0.3 | -0.29 | -0.04 | -0.1 |
| carbohydrate transmembrane transport | -0.17 | 0.32 | 0.17 | 0.04 | -0.08 | 0.11 | -0.22 | 0.07 |
| regulation of neuron death | 0.15 | 0.01 | -0.38 | -0.19 | -0.31 | -0.2 | -0.03 | -0.01 |
| mitochondrion organization | -0.19 | -0.06 | 0.12 | 0.23 | -0.05 | 0.02 | -0.15 | 0.24 |
| neurogenesis | -0.12 | 0.09 | **-0.54*** | 0.32 | -0.13 | -0.05 | -0.12 | 0.25 |
| reproductive behavior | -0.18 | -0.02 | -0.35 | 0.2 | -0.05 | -0.07 | -0.17 | 0.11 |
| Genes not belonging in any other module | 0.05 | 0.24 | 0.36 | 0.36 | 0.06 | 0.26 | 0.07 | 0.07 |
| **Selected gene markers of MARCO+ macrophage subset** | | | | | | | | |
| **Gene** |  | | | | | | | |
| STAB1 | -0.01 | 0.09 | 0.23 | -0.17 | 014 | 0.04 | -0.11 | -0.06 |
| MARCO | -0.23 | -0.03 | **0.48** | -0.05 | 0.16 | 0.07 | -0.1 | -0.07 |
| CD163 | -0.05 | 0.05 | 0.19 | -0.35 | 0.18 | 0 | -0.12 | -0.15 |

Nominally significant correlations (p value < 0.05) are in bold, FDR adjusted significant (FDR <0.05) are denoted by a star*

**Supplemental Table 4: Correlations between MBP or β-tubulin III (immunohistochemistry) and clinical pain phenotype**

| **Clinical pain scores** | **Corrected mean grey values of** | **Correlation test** | **r** | **p-value** | **Adjusted p-value** |
| --- | --- | --- | --- | --- | --- |
| NPSI burning pain | Myelin basic protein | Spearman’s | -0.356 | 0.161 | 0.375 |
| NPSI deep pain | Myelin basic protein | Pearson’s | 0.056 | 0.831 | 0.831 |
| NPSI paroxysmal pain | Myelin basic protein | Spearman’s | 0.080 | 0.759 | 0.886 |
| NPSI evoked pain | Myelin basic protein | Pearson’s | -0.192 | 0.461 | 0.646 |
| NPSI paraesthesia | Myelin basic protein | Spearman’s | -0.391 | 0.122 | 0.427 |
| VAS numbness (24 hours) | Myelin basic protein | Spearman’s | -0.239 | 0.370 | 0.648 |
| VAS pain (24 hours) | Myelin basic protein | Pearson’s | -0.488 | 0.055 | 0.387 |
| NPSI burning pain | β-tubulin III | Spearman’s | -0.162 | 0.531 | 0.743 |
| NPSI deep pain | β-tubulin III | Pearson’s | 0.262 | 0.310 | 0.542 |
| NPSI paroxysmal pain | β-tubulin III | Spearman’s | 0.462 | 0.063 | 0.222 |
| NPSI evoked pain | β-tubulin III | Pearson’s | -0.046 | 0.862 | 1.000 |
| NPSI paraesthesia | β-tubulin III | Spearman’s | -0.271 | 0.292 | 0.680 |
| VAS numbness (24 hours) | β-tubulin III | Spearman’s | -0.022 | 0.936 | 0.936 |
| **VAS pain (24 hours)** | **β-tubulin III** | **Pearson’s** | **-0.653** | **0.006** | **0.043*** |
| **Clinical pain scores** | **Area fractions of** | **Correlation test** | **r** | **p-value** | **Adjusted p-value** |
| NPSI burning pain | Myelin basic protein | Spearman’s | -0.269 | 0.294 | 0.515 |
| NPSI deep pain | Myelin basic protein | Pearson’s | 0.106 | 0.685 | 0.685 |
| NPSI paroxysmal pain | Myelin basic protein | Spearman’s | -0.167 | 0.518 | 0.605 |
| NPSI evoked pain | Myelin basic protein | Pearson’s | -0.212 | 0.414 | 0.580 |
| NPSI paraesthesia | Myelin basic protein | Spearman’s | -0.429 | 0.087 | 0.204 |
| VAS numbness (24 hours) | Myelin basic protein | Spearman’s | -0.469 | 0.069 | 0.241 |
| VAS pain (24 hours) | Myelin basic protein | Pearson’s | -0.552 | 0.027* | 0.186 |
| NPSI burning pain | β-tubulin III | Spearman’s | -0.174 | 0.502 | 0.753 |
| NPSI deep pain | β-tubulin III | Pearson’s | 0.188 | 0.469 | 1.000 |
| NPSI paroxysmal pain | β-tubulin III | Spearman’s | 0.160 | 0.537 | 0.645 |
| NPSI evoked pain | β-tubulin III | Pearson’s | -0.071 | 0.786 | 0.786 |
| NPSI paraesthesia | β-tubulin III | Spearman’s | -0.421 | 0.094 | 0.562 |
| VAS numbness (24 hours) | β-tubulin III | Spearman’s | -0.188 | 0.482 | 0.963 |
| **VAS pain (24 hours)** | **β-tubulin III** | **Pearson’s** | **-0.7096** | **0.002** | **0.015*** |

Choice of correlation tests was based on normality of data. Nominally significant correlations are in bold, FDR adjusted significant p-values are denoted by a star*

**Supplemental Table 5: Correlations between intraneural CD3^+^ or CD68^+^ cells (immunohistochemistry) and clinical pain phenotype**

| **Clinical measurement scores** | **The densities of** | **Correlation test** | **r** | **p-value** | **Adjusted p-value** |
| --- | --- | --- | --- | --- | --- |
| NPSI burning pain | CD3^+^ cells | Spearman’s | 0.466 | 0.061 | 0.366 |
| NPSI deep pain | CD3^+^ cells | Spearman’s | 0.126 | 0.629 | 0.629 |
| NPSI paroxysmal pain | CD3^+^ cells | Spearman’s | 0.166 | 0.521 | 0.626 |
| NPSI evoked pain | CD3^+^ cells | Spearman’s | 0.233 | 0.365 | 0.547 |
| NPSI paraesthesia | CD3^+^ cells | Spearman’s | 0.414 | 0.100 | 0.300 |
| VAS pain (24 hours) | CD3^+^ cells | Spearman’s | 0.297 | 0.261 | 0.523 |
| **NPSI burning pain** | **CD68^+^ cells** | **Spearman’s** | **0.633** | **0.008** | **0.046*** |
| NPSI deep pain | CD68^+^ cells | Pearson’s | 0.142 | 0.586 | 0.703 |
| NPSI paroxysmal pain | CD68^+^ cells | Spearman’s | 0.151 | 0.560 | 0.840 |
| NPSI evoked pain | CD68^+^ cells | Pearson’s | 0.074 | 0.779 | 0.779 |
| NPSI paraesthesia | CD68^+^ cells | Spearman’s | 0.388 | 0.125 | 0.374 |
| VAS pain (24 hours) | CD68^+^ cells | Pearson’s | 0.221 | 0.411 | 0.822 |

Choice of correlation tests was based on normality of data. Nominally significant correlations are in bold, FDR adjusted significant p-values are denoted by a star*

**Supplemental Table 6: Correlations between intraneural CD163^+^ or MARCO^+^ cells (immunohistochemistry) and clinical pain phenotype**

| **Clinical measurement scores** | **The densities of** | **Correlation test** | **r** | **p-value** | **Adjusted p-value** |
| --- | --- | --- | --- | --- | --- |
| NPSI burning pain | CD163^+^ cells | Spearman’s | -0.193 | 0.444 | 0.665 |
| NPSI deep pain | CD163^+^ cells | Spearman’s | -0.019 | 0.941 | 1.000 |
| NPSI paroxysmal pain | CD163^+^ cells | Spearman’s | 0.321 | 0.194 | 1.000 |
| NPSI evoked pain | CD163^+^ cells | Spearman’s | 0.196 | 0.435 | 0.870 |
| NPSI paraesthesia | CD163^+^ cells | Spearman’s | 0.263 | 0.291 | 0.874 |
| VAS pain (24 hours) | CD163^+^ cells | Spearman’s | 0.012 | 0.996 | 0.996 |
| NPSI burning pain | MARCO^+^ cells | Spearman’s | -0.295 | 0.234 | 0.702 |
| NPSI deep pain | MARCO^+^ cells | Spearman’s | 0.192 | 0.445 | 0.668 |
| NPSI paroxysmal pain | MARCO^+^ cells | Spearman’s | 0.312 | 0.207 | 1.000 |
| NPSI evoked pain | MARCO^+^ cells | Spearman’s | 0.139 | 0.583 | 0.699 |
| NPSI paraesthesia | MARCO^+^ cells | Spearman’s | 0.295 | 0.235 | 0.471 |
| VAS pain (24 hours) | MARCO^+^ cells | Spearman’s | -0.081 | 0.758 | 0.758 |

Choice of correlation tests was based on normality of data

**Supplemental Table 7: Correlations between intraneural CD163^+^MARCO^+^ or CD163^+^MARCO^-^ cells (immunohistochemistry) and clinical pain phenotype**

| **Clinical measurement scores** | **The densities of** | **Correlation test** | **r** | **p-value** | **Adjusted p-value** |
| --- | --- | --- | --- | --- | --- |
| NPSI burning pain | CD163^+^MARCO^+^ cells | Spearman’s | -0.211 | 0.402 | 1.000 |
| NPSI deep pain | CD163^+^MARCO^+^ cells | Spearman’s | 0.139 | 0.582 | 0.873 |
| NPSI paroxysmal pain | CD163^+^MARCO^+^ cells | Spearman’s | 0.204 | 0.418 | 1.000 |
| NPSI evoked pain | CD163^+^MARCO^+^ cells | Spearman’s | 0.089 | 0.726 | 0.726 |
| NPSI paraesthesia | CD163^+^MARCO^+^ cells | Spearman’s | 0.193 | 0.444 | 0.888 |
| VAS pain (24 hours) | CD163^+^MARCO^+^ cells | Spearman’s | -0.108 | 0.679 | 0.815 |
| NPSI burning pain | CD163^+^MARCO^-^ cells | Spearman’s | -0.138 | 0.584 | 0.877 |
| NPSI deep pain | CD163^+^MARCO^-^ cells | Pearson’s | 0.035 | 0.890 | 0.890 |
| NPSI paroxysmal pain | CD163^+^MARCO^-^ cells | Spearman’s | 0.424 | 0.079 | 0.476 |
| NPSI evoked pain | CD163^+^MARCO^-^ cells | Pearson’s | -0.061 | 0.810 | 0.972 |
| NPSI paraesthesia | CD163^+^MARCO^-^ cells | Spearman’s | 0.272 | 0.275 | 0.826 |
| VAS pain (24 hours) | CD163^+^MARCO^-^ cells | Pearson’s | -0.191 | 0.462 | 0.923 |

Choice of correlation tests was based on normality of data

**Supplemental Table 8: Correlations between intraneural CD3^+^CD4^+^ cells (immunohistochemistry) and clinical pain phenotype**

| **Clinical measurement scores** | **The densities of** | **Correlation test** | **r** | **p-value** | **Adjusted p-value** |
| --- | --- | --- | --- | --- | --- |
| NPSI burning pain | CD3^+^CD4^+^ cells | Spearman’s | 0.075 | 0.755 | 1.000 |
| NPSI deep pain | CD3^+^CD4^+^ cells | Pearson’s | 0.065 | 0.785 | 1.000 |
| NPSI paroxysmal pain | CD3^+^CD4^+^ cells | Spearman’s | 0.450 | **0.047** | 0.279 |
| NPSI evoked pain | CD3^+^CD4^+^ cells | Pearson’s | 0.018 | 0.940 | 1.000 |
| NPSI paraesthesia | CD3^+^CD4^+^ cells | Spearman’s | 0.077 | 0.747 | 1.000 |
| VAS pain (24 hours) | CD3^+^CD4^+^ cells | Pearson’s | -0.004 | 0.988 | 0.988 |

Choice of correlation tests was based on normality of data. Nominally significant correlations are in bold.

**Supplemental Table 9: Correlations between intraneural MBP or β-tubulin III and immune cells (immunohistochemistry)**

| **Area fraction of** | **The densities of** | **Correlation test** | **r** | **p-value** | **Adjusted p-value** |
| --- | --- | --- | --- | --- | --- |
| **Myelin basic protein** | **CD3^+^ cells** | **Spearman’s** | **-0.6594** | **0.003** | **0.020*** |
| **Myelin basic protein** | **CD68^+^ cells** | **Pearson’s** | **-0.6036** | **0.008** | **0.028*** |
| Myelin basic protein | CD3^+^CD4^+^ cells | Pearson’s | -0.2268 | 0.366 | 0.426 |
| Myelin basic protein | CD163^+^ cells | Spearman’s | -0.3260 | 0.201 | 0.352 |
| Myelin basic protein | MARCO^+^ cells | Spearman’s | 0.3211 | 0.209 | 0.292 |
| Myelin basic protein | CD163^+^MARCO^+^ cells | Spearman’s | 0.0956 | 0.716 | 0.716 |
| Myelin basic protein | CD163^+^MARCO^-^ cells | Pearson’s | -0.4267 | 0.088 | 0.204 |
| β-tubulin III | CD3+ cells | Spearman’s | -0.0341 | 0.893 | 1.000 |
| β-tubulin III | CD68+ cells | Pearson’s | -0.0546 | 0.830 | 1.000 |
| β-tubulin III | CD3+CD4+ cells | Pearson’s | 0.2162 | 0.389 | 0.907 |
| β-tubulin III | CD163^+^ cells | Spearman’s | 0.0808 | 0.7584 | 1.000 |
| β-tubulin III | MARCO^+^ cells | Spearman’s | 0.2598 | 0.3127 | 1.000 |
| β-tubulin III | CD163^+^MARCO^+^ cells | Spearman’s | 0.4657 | 0.0615 | 0.431 |
| β-tubulin III | CD163^+^MARCO^-^ cells | Pearson’s | -0.0114 | 0.965 | 0.965 |

Choice of correlation tests was based on normality of data. Nominally significant correlations are in bold, FDR adjusted significant p-values are denoted by a star*

**Supplemental Figure 1: Location of Morton’s neuroma sample collection.**

Two samples (pink) were taken just proximal to the bifurcation of the plantar digital nerve and used for molecular and cellular analyses. These samples represent the location of the Morton’s neuroma under the transverse metatarsal plantar ligament. The distal branches (green) were stored but not used for analyses in this paper.

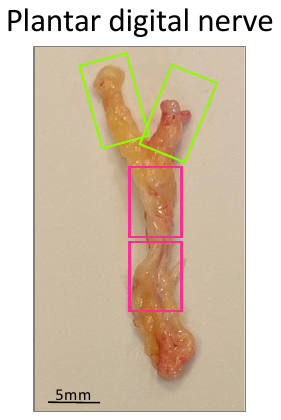

distal

**Supplemental Figure 2: The densities of intraneural CD3^+^CD4^+^ cells are comparable between Morton’s neuroma and healthy control nerves.**

**(A)** Representative images of nerve sections from a patient with Morton’s neuroma (left) and a healthy individual stained with anti-CD3 (red), CD4 (green) antibodies and DAPI (blue). The boundaries of intraneural areas are indicated with white dotted lines. The areas within yellow squares were enlarged 11.5 times and displayed below. The red, green and yellow arrows indicate examples of CD3^+^, CD4+ and CD3^+^CD4^+^ cells respectively. **(B)** The density of intraneural CD3^+^CD4^+^ was comparable between groups. Morton’s Neuroma, N=21; Control Nerve, N=11. Data are presented as means and standard deviations. Student’s t tests, ns: p =0.7147.

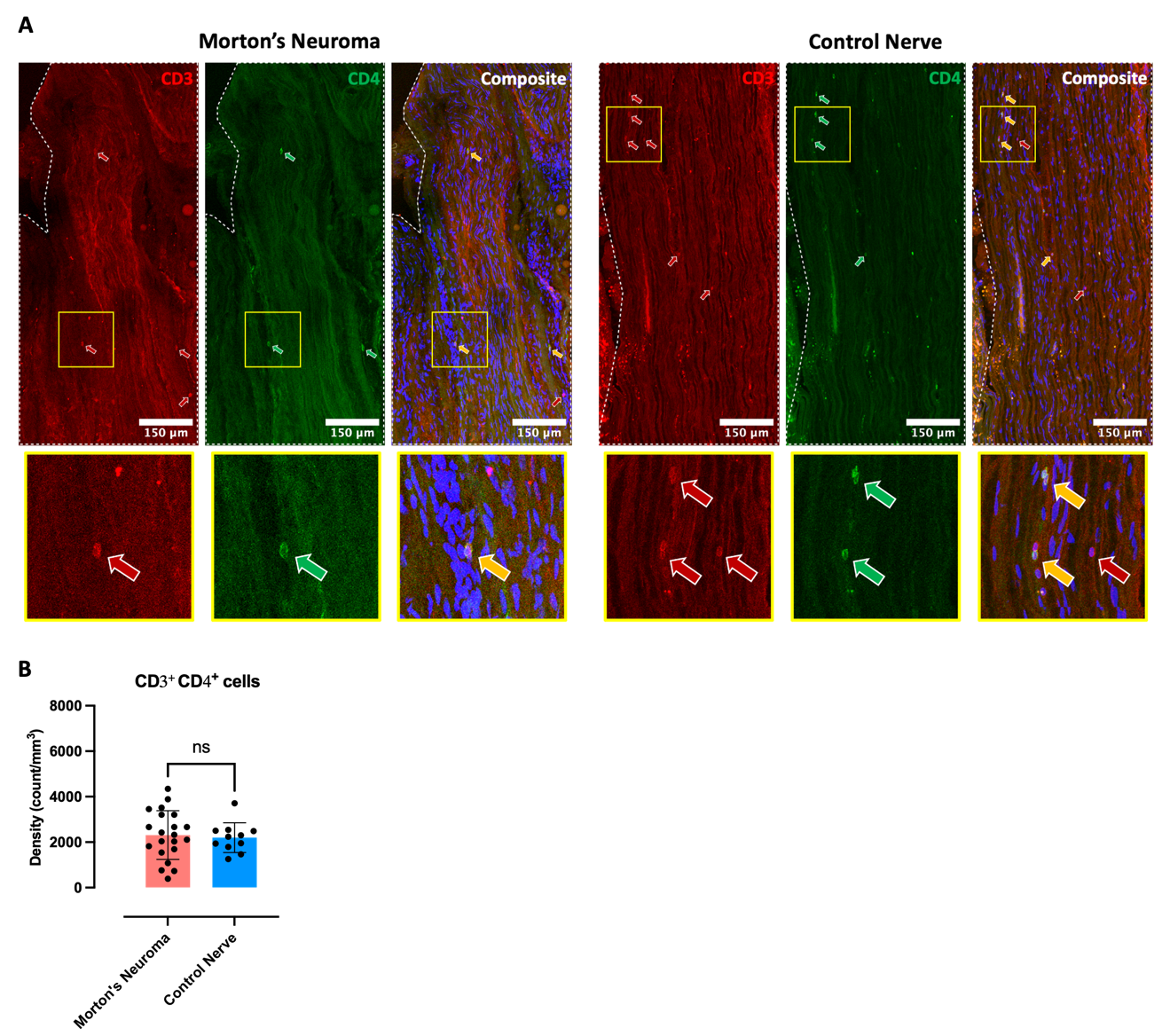

**Supplemental Figure 3: The area fractions of intraneural MBP significantly correlate with the densities of intraneural CD3^+^ and CD68^+^ cells.**

Correlations of area fractions of intraneural MBP with densities of **(A)** intraneural CD3^+^ and **(B)** CD68 cells. The type of correlation test based on normality of data, r and FDR-adjusted p-value are indicated in each plot. N=18, *: p < 0.05

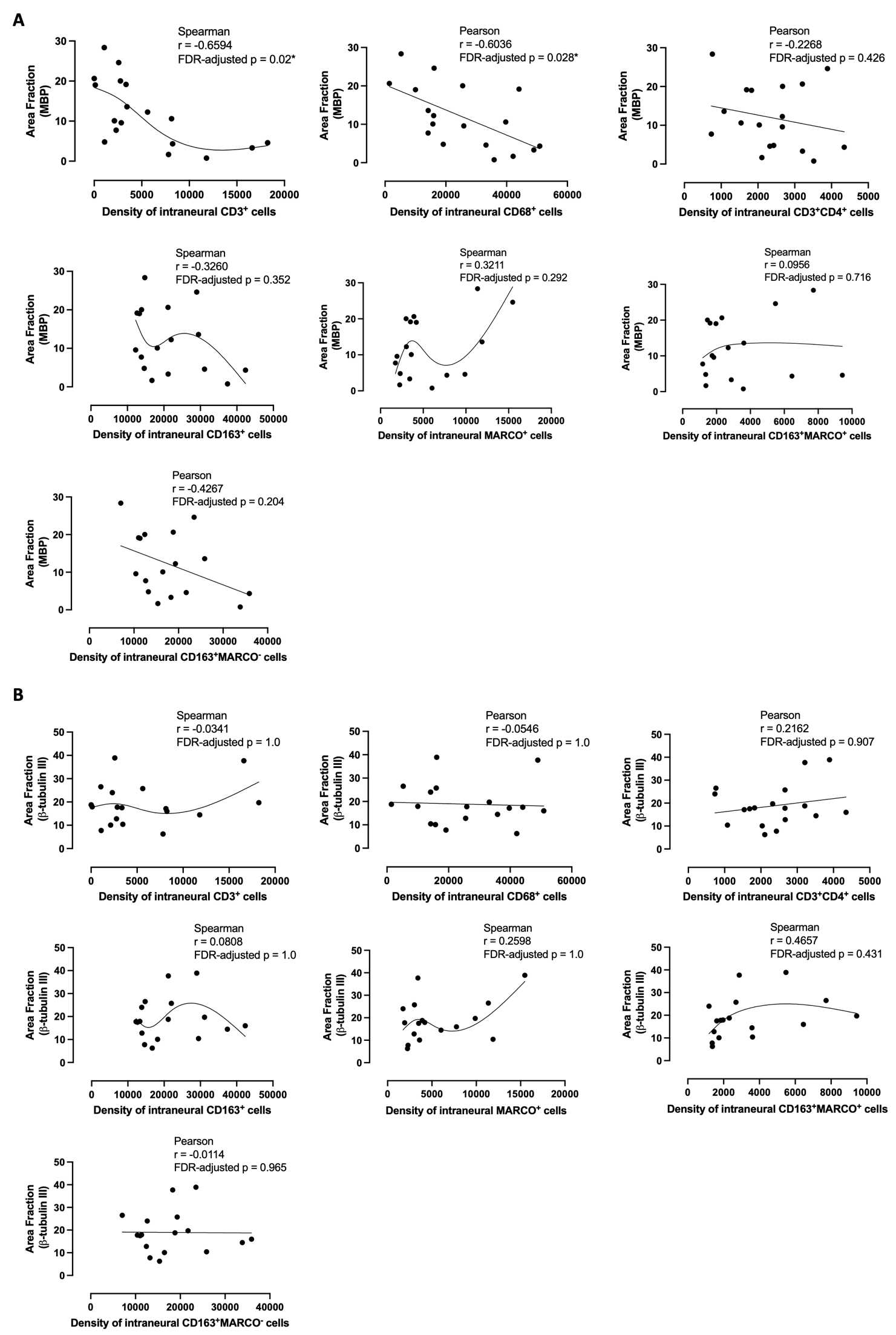

**B**
